# Using expert-cited features to detect leg dystonia in cerebral palsy

**DOI:** 10.1101/2024.10.15.24315574

**Authors:** Rishabh Bajpai, Alyssa Rust, Emma Lott, Susie Kim, Sushma Gandham, Keerthana Chintalapati, Joanna Blackburn, Rose Gelineau-Morel, Michael C. Kruer, Dararat Mingbunjerdsuk, Jennifer O’Malley, Laura Tochen, Jeff L. Waugh, Steve Wu, Timothy Feyma, Joel S. Perlmutter, Bhooma R. Aravamuthan

**Author notes:** Corresponding author. Division of Pediatric Neurology, Department of Neurology Washington University School of Medicine 660 South Euclid Avenue, Campus Box 8111 St. Louis MO 63110-1093.

## Abstract

**Objectives:** Leg dystonia in cerebral palsy (CP) is debilitating but remains underdiagnosed. Routine clinical evaluation has only 12% accuracy for leg dystonia diagnosis compared to gold-standard expert consensus assessment. We determined whether expert-cited leg dystonia features could be quantified to train machine learning (ML) models to detect leg dystonia in videos of children with CP.

**Methods:** Eight pediatric movement disorders physicians assessed 298 videos of children with CP performing a seated task at two CP centers. We extracted leg dystonia features cited by these experts during consensus-building discussions, quantified these features in videos, used these quantifications to train 4664 ML models on 163 videos from one center, and tested the best performing models on a separate set of 135 videos from both centers.

**Results:** We identified 69 quantifiable features corresponding to 12 expert-cited leg dystonia features. ML models trained using these quantifications achieved 88% sensitivity, 74% specificity, 82% positive predictive value, 84% negative predictive value, and 82% accuracy for identifying leg dystonia across both centers. Of the 25 features contributing to the best performing ML models, 17 (68%) quantified leg movement variability. We used these ML models to develop DxTonia, open-source software that identifies leg dystonia in videos of children with CP.

**Interpretation:** DxTonia primarily leverages detection of leg movement variability to achieve 82% accuracy in identifying leg dystonia in children with CP, a significant improvement over routine clinical diagnostic accuracy of 12%. Observing or quantifying leg movement variability during a seated task can facilitate leg dystonia detection in CP.

## Introduction

Dystonia is a debilitating movement disorder characterized by abnormal movements worsened by voluntary action and associated with overflow.^1^ Cerebral palsy (CP) is the most common childhood cause of dystonia, affecting 2–4 of every 1000 people.^2–4^ Differentiating dystonia from other co-occurring movement conditions in CP, like spasticity, is crucial because dystonia and spasticity treatments differ.^5,6^ Dystonia can also contraindicate some surgeries targeting spasticity in children with CP.^7–9^ For example, seemingly mild leg dystonia may be “unmasked” following selective dorsal rhizotomy if not detected in advance.^7,10^ Therefore, identification of even mild leg dystonia can be critical for treatment selection.

Dystonia is variable,^1,11^ making it difficult to diagnose as a single clinician in a busy clinic. Gold standard diagnosis requires consensus from multiple pediatric movement disorder experts based on video or in-person assessments. However, these experts are few and unavailable at many centers.^12^ Routine clinical assessment by a single pediatric movement disorders physician has only 12% accuracy for leg dystonia diagnosis in a person with CP during gait, as compared to gold standard consensus-based expert assessment by those same physicians.^13^ Thus, it is necessary to improve clinical leg dystonia diagnosis in CP.

Currently, the Hypertonia Assessment Tool (HAT) remains the only clinical tool available for distinguishing dystonia from spasticity in CP. However, dystonia-specific HAT items have only moderate interrater reliability and require detection of subtle movements, thus necessitating significant clinician time and expertise.^14,15^

We have shown that experts cite leg adduction variability and amplitude when identifying leg dystonia in people with CP during gait.^16^ We have also shown that quantifications of leg adduction variability and amplitude correlate with expert-assessed leg dystonia severity during gait (when the legs are actively engaged)^17^ and during a seated hand open-close task^18^ (when the legs are not actively engaged, thus representing overflow dystonia^1^ as per the 2025 consensus definition). However, it is unclear whether these features distinguish between the presence and absence of dystonia, which remains a critical question particularly for surgical candidacy.

We hypothesized that machine learning (ML) models trained using expert-cited leg dystonia features could identify leg dystonia with higher accuracy than routine clinical assessment. We chose this approach, as opposed to an unsupervised ML approach, to train models to represent key features of expert assessment and not spurious features only co-segregating with leg dystonia without representing dystonia itself.^19–21^ We additionally hypothesized that the best discriminating features of leg dystonia would be leg adduction variability and amplitude.^17,18^ To test these hypotheses, we had eight pediatric movement disorders physicians establish their consensus assessment for leg dystonia in videos of children with CP performing a seated hand open-close task. We determined the leg dystonia features they cited in their consensus-building discussions and quantified these features in clinically-acquired videos. These quantifications were used to train ML models for leg dystonia diagnosis. Movement features were ranked based on their relative contributions to the ML models that best identified leg dystonia. Finally, we used these ML models to develop DxTonia, open-source software to facilitate automated clinical screening for leg dystonia in people with CP.

## Methods

### Standard protocol approvals and registrations

This study received approval from the Washington University in St. Louis Institutional Review Board on 6/5/23 (Approval Number: 202102101).

### Participants, motor task, and video acquisition

We obtained video recordings of children with CP performing a seated alternating hand open-close task during routine clinical care between 1/1/2020–12/30/2021 at Gillette Children’s Hospital CP Center (St. Paul, MN, USA, henceforth called Center 1) and between 4/20/2023– 12/14/2023 at St. Louis Children’s Hospital CP Center (St. Louis, MO, USA, henceforth called Center 2). The task involves resting one hand on the lap, while raising the other hand to eye level and opening and closing that hand as quickly as possible for 3-5 seconds. The raised hand is then placed on the lap, and the other hand is raised and opened/closed. A single task cycle involves opening/closing one hand and then the other. At Center 1, videos contained only one task cycle recorded with a Panasonic AG-AC160AP camera at 1920 x 1080 pixel resolution at 30 frames per second. At Center 2, videos contained five task cycles recorded with a Google Pixel 5 smartphone camera at 3840 x 2160 pixel resolution at 60 frames per second rate. We included potential participants in this study if they (**1**) were aged five years or older, to increase the likelihood that they had the cognitive ability and attention span to attempt the motor task in full, and (**2**) had a CP Center clinician-confirmed diagnosis of CP, as per the widely-used 2006 definition of CP.^22^ We excluded potential participants if (**1**) their full body was not visible in the video recording, or (**2**) they were unable to complete the task during the video recording. Clinical characteristics of the subjects were determined via prospective data entry by the clinical providers at both Centers (including Gross Motor Function Classification System level and presence of spasticity as determined using the Modified Ashworth Scale at Center 1 and the Modified Tardieu Scale at Center 2). The detailed protocol can be found in eAppendix 1.

Use of the seated alternating hand open-close task for dystonia assessment is valuable for numerous reasons. First, our previous comparison of leg dystonia severity ratings during seated upper extremity tasks, gait, and a comprehensive 30-minute exam protocol suggested that seated upper extremity tasks may best approximate leg dystonia severity ratings compared to the comprehensive exam protocol.^23^ Second, we have previously shown that leg adduction variability and amplitude metrics track with experts’ leg dystonia severity ratings during the seated alternating hand open-close task,^18^ suggesting that quantifiable metrics may be valuable for discriminating leg dystonia during this task. Third, given that variability is a well-established feature of dystonia, distinguishing movement variability due to dystonia and movement variability due to other causes (including voluntary movement) is easiest in a task where the legs are otherwise supposed to be still, as they would be in a seated upper extremity task vs. gait. Finally, not all people with dystonia and CP can walk. Using a seated task, as opposed to gait, facilitates broader access to any tools developed for dystonia assessment.

Video recordings of the seated alternating hand open-close task show participants seated in a chair facing forward resting their non-dominant hand on the ipsilateral thigh, and then raising and alternately opening and closing/fisting their dominant hand as quickly as possible for approximately five seconds. The task is then repeated with the non-dominant hand. Noting that this task should require only upper limb voluntary movement, overflow dystonia in the legs could occur as outlined in the 2025 consensus dystonia definition which describes voluntary movement in an unaffected part of the body causing overflow dystonic movements and postures into apparently unaffected body segments.^1^

### Consensus-based expert video assessment

To ensure participant anonymity, we blurred the faces of all participants in recordings using ShotCut (Meltytech, LLC) and custom-written software.^24^ To achieve consensus, eight pediatric movement disorders physicians with CP expertise practicing at eight different institutions across the United States (J.B., R.G.M., M.K., D.M., J.O.M., L.T., J.W., S.W.) rated all videos after participating in six one-hour Zoom sessions (Zoom Video Communications, Inc.) to discuss leg dystonia severity in a consensus building dataset of 54 videos (24 from Center 1 and 30 from Center 2). Notably, none of these eight experts were from Centers 1 or 2. The initial 54 videos were selected as follows:

Center 1: A pediatric movement disorders physician with expertise in CP (B.R.A.) assessed all 193 videos for leg dystonia using the GDRS. These preliminary ratings, which were never shared with the other expert raters or incorporated into the final ratings, were used only to select 24 videos that were putatively representative of the full range of leg dystonia severities in the full 193 video dataset. These 24 videos were split into three groups of 8 videos (also representing the full range of dystonia severities) which were discussed by all the other expert raters across three consensus-building Zoom sessions.

Center 2: Using our published standardized and prospective clinical data entry workflow,^25^ 30 videos of the 105 were selected to represent a range of GMFCS levels (I-III) and dystonia as the predominant tone type, accompanying tone type, or absent tone type. Again, these clinician determinations were not shared with the expert raters or incorporated into the final ratings. These 30 videos were split into three groups of 10 (also representative of the above ranges of GMFCS and tone classifications) and were discussed by all the other expert raters across three consensus-building Zoom sessions.

After these consensus-building sessions, experts rated the remaining 244 videos from both Centers and recorded scores via a REDCap survey. We calculated the final assessment of leg dystonia severity for each video as the median Global Dystonia Rating Scale (GDRS),^26^ across all expert reviewers, with GDRS < 1 indicating leg dystonia was absent and GDRS ≥ 1 indicating that leg dystonia was present.

### Qualitative analysis of consensus-building discussions

We used conventional content analysis^27^ to analyze transcripts of expert consensus-building discussions to identify the salient ideas, or codes, stated by experts when assessing videos for leg dystonia. Two coders (SG, KC), independently coded the transcripts, met to resolve any coding discrepancies by consensus, and then consolidated their codes into a single code book. We distinguished potentially quantifiable movements (e.g. “foot inversion”) from non-quantifiable codes (e.g. “posturing”) and used the quantifiable movements to calculate kinematics analogues from videos for training ML models.

### Extraction of leg coordinates

We used custom-written software^24^ that leverages OpenPose^28^ to estimate the following consistently visualizable leg coordinates from the videos: the midpoint of the patella, the midpoint between the medial and lateral malleolus, and the tip of the first toe for both the right and left legs. To ensure labeling accuracy, we visually examined the extracted coordinates overlayed across all video frames for a subset of videos. Coordinates were normalized to correct for patient height (by dividing by the maximum knee-to-ankle distance^18^) and camera positioning (by subtracting the median pelvis coordinates).

### Determining the features used to train dystonia diagnostic models

We calculated leg kinematics based on: (**1**) 2D leg coordinates (n = 12, X and Y coordinates of the right and left knee, ankle, and first toe), and (**2**) analogues of quantifiable movements cited by experts when assessing videos for dystonia. Since we previously demonstrated that variability and amplitude of leg adduction are cited by experts when assessing leg dystonia severity and also correlate with their GDRS-based assessments of leg dystonia severity,^17^ we used the variance, minimum, and maximum of all leg kinematics measures as the input features to train ML models for leg dystonia diagnosis.

### Dystonia diagnostic model training, validation, and testing

We evaluated a total of 4664 ML models derived from 11 different ML algorithms: Logistic Regression,^29^ Decision Tree,^30^ Random Forest,^31^ Gradient Boosting,^32^ AdaBoost,^33^ Support Vector Machine,^34^ K-Nearest Neighbors,^35^ Naive Bayes,^36^ XGBoost,^37^ Extra Trees,^38^ and Bagging.^39^ These 4,664 ML models have different learning parameters and algorithms, and therefore exhibit different learning capabilities. We divided videos from Center 1 into three subsets for training (2/3 of the dataset), validation (1/6 of the dataset), and testing (1/6 of the dataset), while ensuring that each subset had comparable GDRS score distributions. To avoid bias from any single arbitrary division into subsets, we re-grouped the full video dataset into different subsets five times and trained, validated, and tested ML models on these five different subset groupings. We repeated training and validation five times within each grouping using different subsets of videos for training and validation each time (five-fold cross-validation using the scikit-learn library) (**Fig 1**).

**Figure 1.**
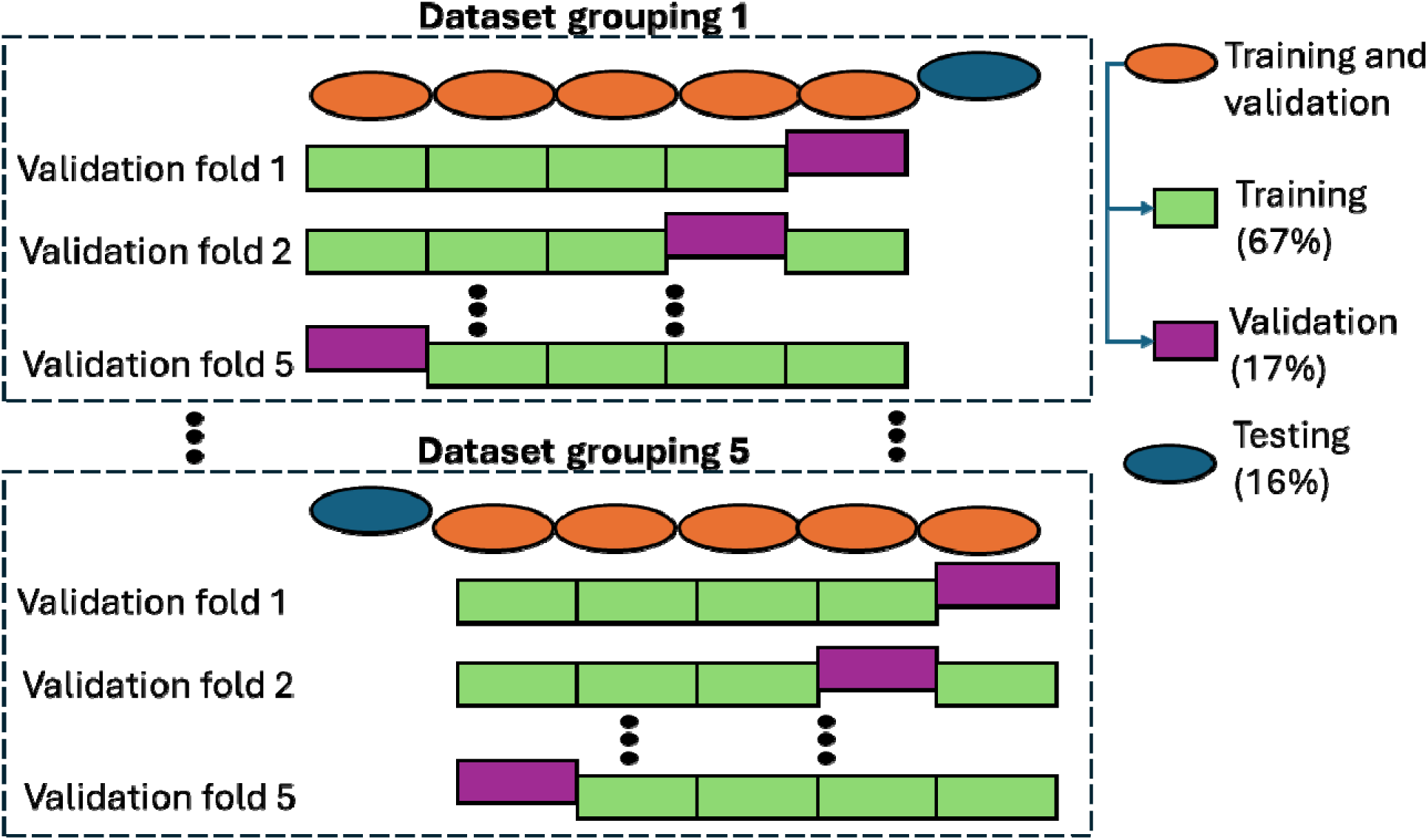
Training, validation, and testing of machine-learning diagnostic model for leg dystonia. We divided the full video dataset five times into different subsets for training (2/3 of the dataset), validation (1/6 of the dataset), and testing (1/6 of the dataset) to generate five different dataset groupings. We then conducted training and validation five times within each of the dataset groupings, using different subsets for training and validation each time.

Of the 4664 ML models used for training and validation for each dataset grouping, we selected the ML model with the highest average negative predictive value (NPV) and specificity across all five validation folds. To determine the optimal number of ML model features needed to maximize NPV and specificity, we first calculated feature importance within each dataset grouping using eight feature selection methods: Spearman’s rho,^40^ Kendall’s tau,^41^ ANOVA F-test/univariate feature selection,^42^ Extra Trees Classifier,^38^ Random Forest,^31^ Maximum Relevance - Minimum Redundancy,^43^ Sequential Feature Selection (forward),^44^ and Recursive Feature Elimination.^45^ We then ranked features in descending order of feature importance for each grouping averaged across the eight feature selection methods, from 1 to N. We first trained models on just the top ranked feature, then the top two ranked features, then the top three ranked features, and so on, until the model was trained using all features. We then tested the model trained on the optimal number of features to maximize NPV and specificity using the testing dataset from each of the five dataset groupings to determine the sensitivity, specificity, positive predictive value (PPV), NPV, and accuracy of the model for leg dystonia diagnosis using videos from Center 1.

Of the N features used to train the ML models, we determined the most relevant features for leg dystonia diagnosis in two ways, (**1**) by comparing the distributions of feature importance rankings across variance, minimum, and maximum features using a one-way ANOVA, and (**2**) by identifying training features shared by the ML models that maximized NPV and specificity across all five dataset groupings.

We created an average ensemble of the ML models maximizing NPV and specificity from each of the five dataset groupings from Center 1 and tested this average ensemble using the separate dataset of videos from Center 2. Ensemble averaging is a common method used to aggregate the results of multiple ML models and is the simple average of each model’s output.^46^ Here, we averaged the dichotomous results of each of the five models for each participant (0 = leg dystonia absent, 1 = leg dystonia present) and rounded to the nearest integer value to determine the average ensemble determination.

### Software graphical user interface development

We used the average ensemble ML model described above plus 250 pre-defined and 2 custom libraries (OpenPose^28^ and exiftool^47^, both open source) to develop open-source software we call DxTonia^24^ (**Supplementary eAppendix 2**). To maximize performance and generalizability, the final uploaded models were re-trained on the combined dataset from both centers. We designed DxTonia to provide an interactive graphical user interface (GUI) to automate leg dystonia detection in user-uploaded videos of children doing a seated alternating hand open/close task. We chose to make DxTonia available for free download and use without requiring installation. Methodologic details regarding the architecture of the DxTonia software package are in Supplementary eFigure 2.

**Figure 2.**
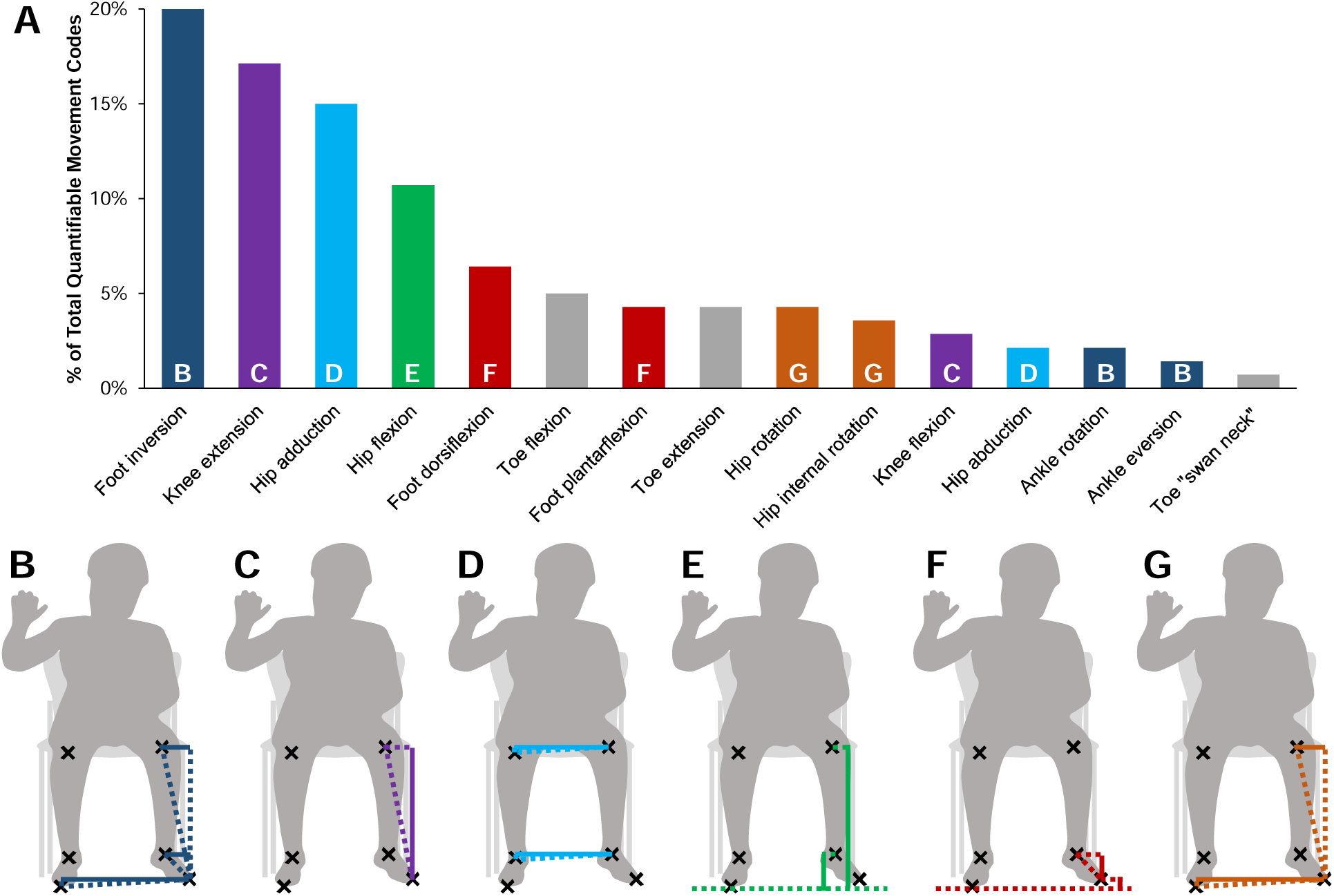
Quantifiable movements cited by experts when diagnosing dystonia. **A)** Frequency with which each quantifiable movement was cited by experts, color coded into groups based on how each movement could be quantified. **B-G)** Example kinematic analogues of expert-cited quantifiable movements (see Tables 1 and 2).

**Table 1.**
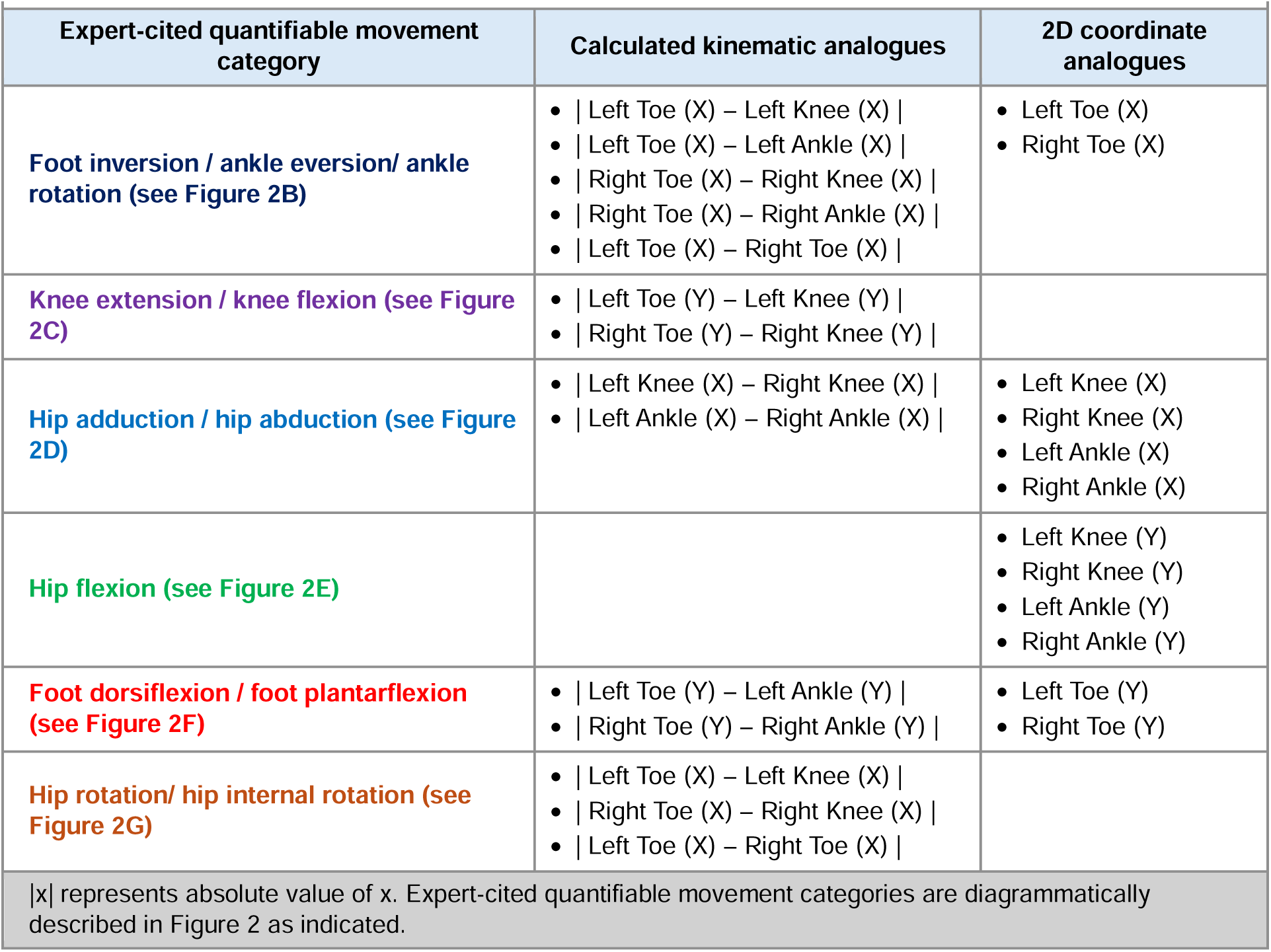
Expert cited quantifiable movements, calculated analogues, and 2D coordinate analogues.

| Expert-cited quantifiable movement category | Calculated kinematic analogues | 2D coordinate analogues |
| --- | --- | --- |
| **Foot inversion / ankle eversion/ ankle rotation (see Figure 2B)** | • \| Left Toe (X) – Left Knee (X) \|<br>• \| Left Toe (X) – Left Ankle (X) \|<br>• \| Right Toe (X) – Right Knee (X) \|<br>• \| Right Toe (X) – Right Ankle (X) \|<br>• \| Left Toe (X) – Right Toe (X) \| | • Left Toe (X)<br>• Right Toe (X) |
| **Knee extension / knee flexion (see Figure 2C)** | • \| Left Toe (Y) – Left Knee (Y) \|<br>• \| Right Toe (Y) – Right Knee (Y) \| |  |
| **Hip adduction / hip abduction (see Figure 2D)** | • \| Left Knee (X) – Right Knee (X) \|<br>• \| Left Ankle (X) – Right Ankle (X) \| | • Left Knee (X)<br>• Right Knee (X)<br>• Left Ankle (X)<br>• Right Ankle (X) |
| **Hip flexion (see Figure 2E)** |  | • Left Knee (Y)<br>• Right Knee (Y)<br>• Left Ankle (Y)<br>• Right Ankle (Y) |
| **Foot dorsiflexion / foot plantarflexion (see Figure 2F)** | • \| Left Toe (Y) – Left Ankle (Y) \|<br>• \| Right Toe (Y) – Right Ankle (Y) \| | • Left Toe (Y)<br>• Right Toe (Y) |
| **Hip rotation/ hip internal rotation (see Figure 2G)** | • \| Left Toe (X) – Left Knee (X) \|<br>• \| Right Toe (X) – Right Knee (X) \|<br>• \| Left Toe (X) – Right Toe (X) \| |  |
| \|x\| represents absolute value of x. Expert-cited quantifiable movement categories are diagrammatically described in Figure 2 as indicated. |  |  |

**Table 2.** Features used to train all 5 ML models across all 5 dataset groupings.

| Calculated kinematic analogue | Feature importance ranking | Expert-cited quantifiable movement category (see Figure 2, Table 1) |
| --- | --- | --- |
| **Variance Features** |  |  |
| Variance \|Right Toe(X) - Right Ankle (X)\| | 1 | Foot inversion / ankle eversion/ ankle rotation (see Figure 2B) |
| Variance \|Left Toe(X) - Left Ankle (X)\| | 2 | Foot inversion / ankle eversion/ ankle rotation (see Figure 2B) |
| Variance Left Toe (X) | 3 | Foot inversion / ankle eversion/ ankle rotation (see Figure 2B) |
| Variance Left Ankle (Y) | 4 | Hip flexion (see Figure 2E) |
| Variance Right Toe (Y) | 5 | Foot dorsiflexion / foot plantarflexion (see Figure 2F) |
| Variance \|Left Ankle (X) – Right Ankle (X)\| | 6 | Hip adduction / hip abduction (see Figure 2D) |
| Variance Left Toe (Y) | 7 | Foot dorsiflexion / foot plantarflexion (see Figure 2F) |
| Variance \|Left Toe(Y) - Left Ankle (Y)\| | 8 | Foot dorsiflexion / foot plantarflexion (see Figure 2F) |
| Variance \|Left Toe (X) – Right Toe (X)\| | 9 | Foot inversion / ankle eversion/ ankle rotation (see Figure 2B)<br>Hip rotation/ hip internal rotation (see Figure 2G) |
| Variance \|Right Toe(Y) – Right Ankle (Y)\| | 10 | Foot dorsiflexion / foot plantarflexion (see Figure 2F) |
| Variance \|Left Toe(Y) - Left Knee (Y)\| | 11 | Knee extension / knee flexion (see Figure 2C) |
| Variance Right Ankle (Y) | 12 | Hip flexion (see Figure 2E) |
| Variance Right Toe (X) | 13 | Foot inversion / ankle eversion/ ankle rotation (see Figure 2B) |
| Variance \|Right Toe(Y) - Right Knee (Y)\| | 15 | Knee extension / knee flexion (see Figure 2C) |
| Variance \|Left Toe(X) – Left Knee (X)\| | 18 | Foot inversion / ankle eversion/ ankle rotation (see Figure 2B)<br>Hip rotation/ hip internal rotation (see Figure 2G) |
| Variance Right Ankle (X) | 19 | Hip adduction / hip abduction (see Figure 2D) |
| Variance Left Ankle (X) | 20 | Hip adduction / hip abduction (see Figure 2D) |
| **Minimum Features** |  |  |
| Minimum \|Left Toe(Y) - Left Ankle (Y)\| | 14 | Foot dorsiflexion / foot plantarflexion (see Figure 2F) |
| Minimum \|Right Toe(Y) – Right Ankle (Y)\| | 21 | Foot dorsiflexion / foot plantarflexion (see Figure 2F) |
| Minimum \|Left Ankle (X) – Right Ankle (X)\| | 23 | Hip adduction / hip abduction (see Figure 2D) |
| Minimum Left Toe (Y) | 25 | Foot dorsiflexion / foot plantarflexion (see Figure 2F) |
| **Maximum Features** |  |  |
| Maximum \|Left Toe(Y) -Left Ankle (Y)\| | 16 | Foot dorsiflexion / foot plantarflexion (see Figure 2F) |
| Maximum \|Right Toe(Y) – Right Ankle (Y)\| | 17 | **Foot dorsiflexion / foot plantarflexion (see Figure 2F)** |
| Maximum \|Left Toe (X) – Right Toe (X)\| | 22 | **Foot inversion / ankle eversion/ ankle rotation (see Figure 2B)**<br>**Hip rotation/ hip internal rotation (see Figure 2G)** |
| \|x\| represents absolute value of x. Expert-cited quantifiable movement categories are diagrammatically described in Figure 2 as indicated. |  |  |

Running DxTonia as currently designed requires a Windows operating system (Microsoft Corporation, Redmond, WA), a graphical processing unit (GPU), and 8GB of VRAM. We tested DxTonia using three combinations of operating systems and GPUs: Windows 11 pro with NVIDIA GeForce RTX4090 GPU (NVIDIA Corporation, Santa Clara, CA), Windows 11 pro with NVIDIA GeForce RTX3080, and Windows 10 pro with NVIDIA GeForce RTX3080. The GUI is designed in Tkinter^48^ to facilitate future cross-platform use. The backend is written in Python and linked to third-party tools such as OpenPose^28^ and exiftool^47^ using system subprocesses. Model weights and the full source code are available through SourceForge^24^ for developers who want to build the software from the source code. For users who do not want to modify the source code, a Windows executable file (created using PyInstaller)^49^ is also available at SourceForge^24^ and allows for use of DxTonia without installation.

To estimate the installation time for developers to build the software from the source code, we tested installation on systems with the following configurations:

1. Mid-tier consumer computer: IntelR Core™ i7 11800H CPU @ 2.40 GHz 16 cores, 32-GB RAM, NVIDIA GeForce RTX3070 GPU with 8GB memory, 64-bit Windows 11 Pro Operating System.
2. High-tier consumer computer: IntelR Core™ i9 13900K CPU @ 3.00 GHz 24 cores, 196-GB RAM, NVIDIA GeForce RTX4090 GPU with 24GB memory, 64-bit Windows 11 Pro Operating System, and

To determine the video processing time for all automated steps (from initial video resolution correction to the generation of leg dystonia assessment reports as CSV files), we evaluated DxTonia across 8 possible scenarios that varied by:

1. System configuration: High-tier vs. mid-tier consumer computer as described above.
2. Video quality: Low quality (1280×720 pixels, 30 fps) vs. high quality (3840×2160 pixels, 60 fps), and
3. Video upload methodology: Sequential upload of individual videos vs. Batch upload of 10 videos at a time.

## Results

### Participants and demographics

We identified videos of 193 children from Center 1 that met our inclusion and exclusion criteria. The demographics of this sample were age 7.2 ± 3.1 years (mean ± SD), 92.75% at Gross Motor Function Classification System Levels I-III, 91% with co-existing leg spasticity, and had a GDRS range 0-21. Of these participants, 54% (n = 104) had an average GDRS ≥ 1 and were thus designated as having leg dystonia (**Supplementary eTable 1**). Videos from Center 1 were 7.2 ± 3.5 seconds in duration.

We identified videos of 105 children from Center 2 that met our inclusion and exclusion criteria. The demographics of this sample were age 10.9 ± 3.7 years, 100% at Gross Motor Function Classification System Levels I-III, 87% with co-existing leg spasticity, and had a GDRS range 0-14. Of these participants, 60% (n = 63) had leg dystonia, determined by average GDRS ≥ 1 (**Supplementary eTable 1**). Videos from Center 2 were 58.5 ± 5.8 seconds in duration.

### Features used to train ML models

In addition to 2D leg coordinates (n = 12 for X and Y coordinates of the left and right knee, ankle, and first toe), we also calculated kinematic analogues using these 2D leg coordinates to reflect features cited by experts when assessing dystonia.

Expert clinicians cited 15 potentially quantifiable movements a total of 140 times over three hours of consensus building discussion when assessing videos for dystonia (**Fig 2A**). We used these quantifiable movements to develop 11 kinematic analogues (**Fig 2B**, **Table 1**). We did not calculate analogues for three out of fifteen quantifiable movements cited by experts, namely, toe flexion, toe extension and “swan neck” toe posturing, as this would require reliable extraction of the coordinates of multiple toes, which could not be accurately and reliably obtained using OpenPose following our visual quality inspection.

### Comparison of feature importance rankings

The variance, minimum, and maximum of the coordinates (n=12) and calculated kinematic analogues (n=11) yielded 69 features that we used for training ML models. The importance rankings of these features across all five dataset groupings are shown in **Supplementary eTable 2**. We found that variance features ranked significantly higher than minimum and maximum features (one-way ANOVA, p < 0.005), **Fig 3**.

**Figure 3.**
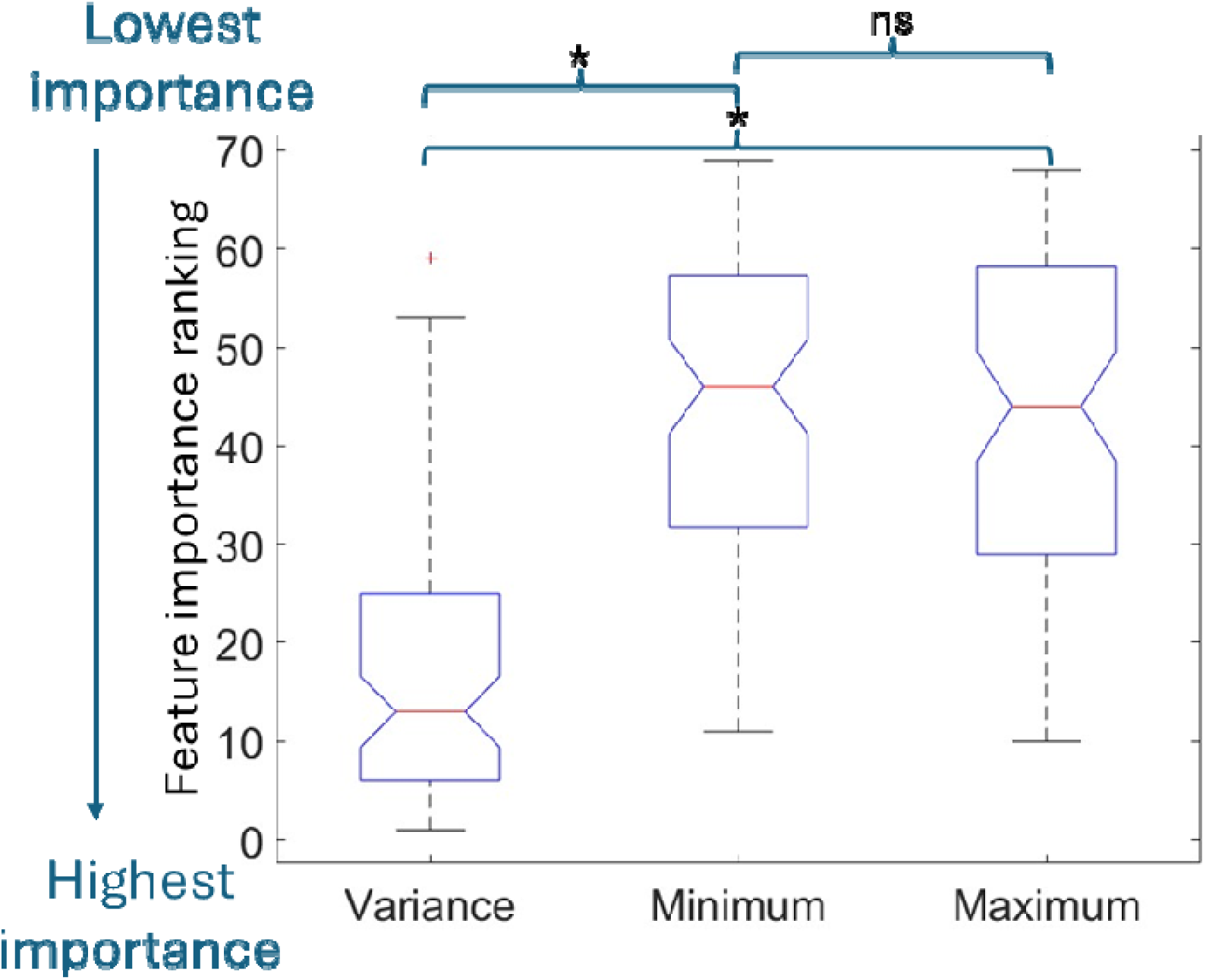
Comparison of the mean importance ranking of variance, minimum, and maximum features. Features were ranked from 1 (highest importance) to 69 (lowest importance). The mean feature importance ranking for variance features was significantly better than the mean rankings for minimum and maximum features (one-way ANOVA, *p < 0.005). Whiskers indicate the interquartile range, the upper and lower edges of the boxes represent the 75th and the 25th percentile of the distribution, respectively, and the red middle line represents the median of the distribution.

### High performing features and diagnostic accuracy of ML models

Of the 4664 ML models based on 11 ML algorithms we evaluated, the models with the highest NPV and specificity were all based on the Naive Bayes^36^ algorithm in all five dataset groupings. These five ML models were trained on the following numbers of top-ranked features: 38, 50, 31, 38, and 35 (**Fig 4**). Of these features, 25 were shared across all five ML models of which 17 were variance features, 4 were minimum features, and 4 were maximum features (**Table 2**). Across all five dataset groupings from Center 1, ML models obtained a mean sensitivity, specificity, PPV, NPV, and accuracy of 87%, 85%, 84%, 88% and 86%, respectively, during validation and 84%, 85%, 85%, 85% and 85%, respectively, during testing (**Table 3**). When tested on 105 videos from Center 2, the average ensemble model had sensitivity, specificity, PPV, NPV, and accuracy of 92%, 62%, 78%, 83%, and 80%, respectively (**Table 3**). Averaged across both centers (noting that leg dystonia prevalence across centers was comparable), the average ensemble model achieves 88% sensitivity, 74% specificity, 82% PPV, 84% NPV, and 82% accuracy compared to gold standard expert consensus diagnosis.

**Figure 4.**
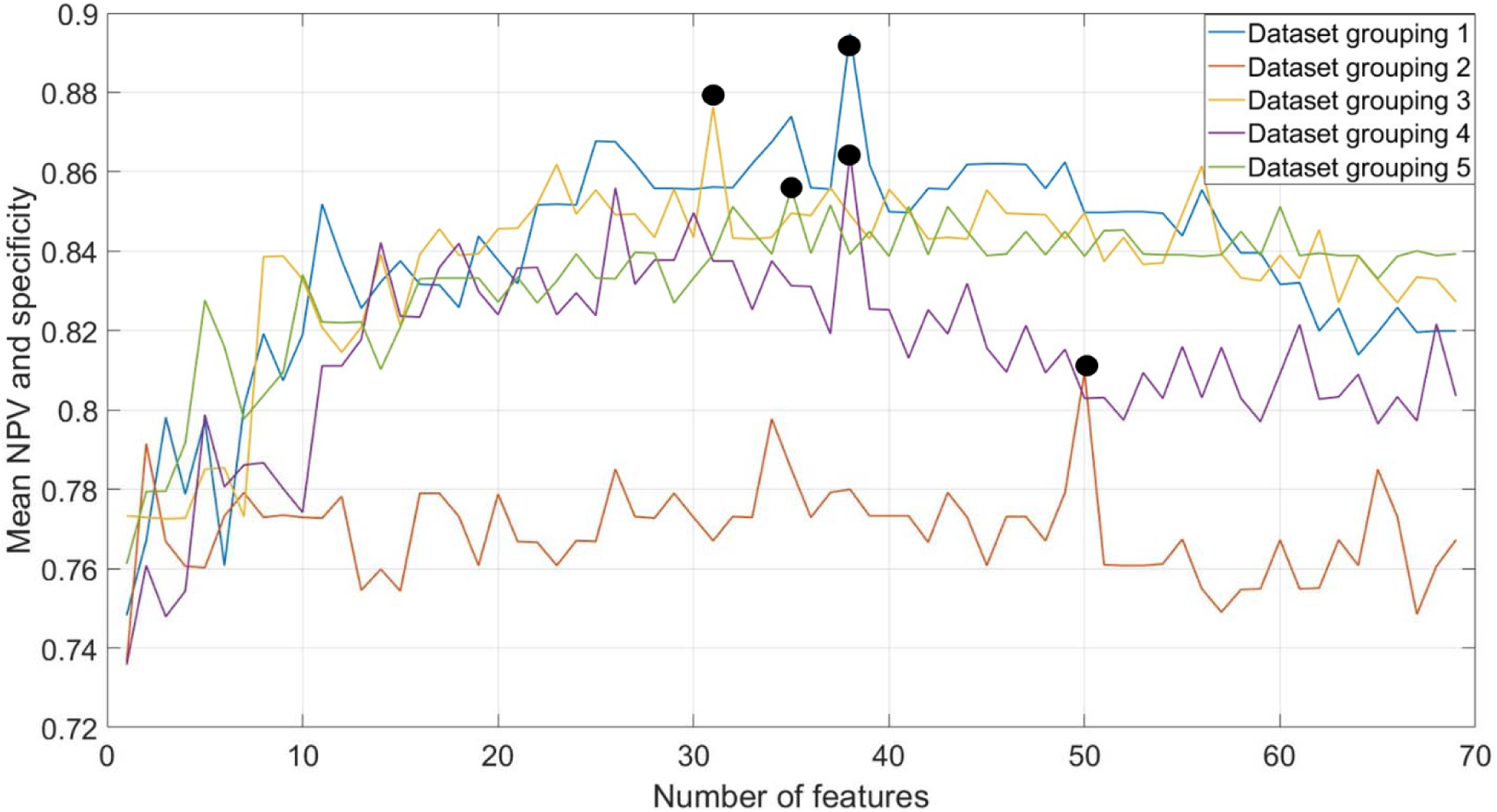
The mean NPV and specificity of ML models when trained on different numbers of top-ranked features. The black points denote the best performing model for each of the five dataset groupings of the full dataset. The x-axis represents the number of features used for training and validation the modles. The y-axis represents the mean validation NPV and specificity of the best-performing ML model (out of 4664 models) when trained on the selected number of features.

**Table 3.** ML model performance during validation and testing across all five dataset groupings from Center 1, and average ensemble model performance on data from Center 2.

**Validation performance of highest performing model from each dataset grouping from Center 1**
| Dataset Grouping | Sensitivity (%) | Specificity (%) | PPV (%) | NPV (%) | Accuracy (%) | Optimal number of features |
| --- | --- | --- | --- | --- | --- | --- |
| 1 | 91 | 86 | 85 | 91 | 88 | 38 |
| 2 | 81 | 81 | 79 | 83 | 82 | 50 |
| 3 | 85 | 91 | 87 | 88 | 88 | 31 |
| 4 | 86 | 86 | 86 | 88 | 86 | 38 |
| 5 | 91 | 80 | 81 | 91 | 85 | 35 |
| Mean | 87 | 85 | 84 | 88 | 86 | 38.4 |

Testing performance of highest performing model from each dataset grouping from Center 1
| Dataset Grouping | Sensitivity (%) | Specificity (%) | PPV (%) | NPV (%) | Accuracy (%) |  |
| --- | --- | --- | --- | --- | --- | --- |
| 1 | 79 | 93 | 93 | 85 | 88 | - |
| 2 | 98 | 81 | 83 | 98 | 89 | - |
| 3 | 95 | 88 | 87 | 92 | 90 | - |
| 4 | 80 | 75 | 77 | 78 | 78 | - |
| 5 | 69 | 86 | 84 | 72 | 79 | - |
| Mean | 84 | 85 | 85 | 85 | 85 | - |

Testing performance from Center 2
| Average Ensemble Model | Sensitivity (%) | Specificity (%) | PPV (%) | NPV(%) | Accuracy (%) |  |
| --- | --- | --- | --- | --- | --- | --- |
| All 105 videos | 92 | 62 | 78 | 83 | 80 | - |

NPV – negative predictive value; PPV- positive predictive value.

### DxTonia installation and video processing times

DxTonia can be used immediately without installation by running the Windows executable file available for free through SourceForge.^24^ For developers who wish to build DxTonia from the source code, installation times vary based on the system configuration. DxTonia installation took 8 minutes 52 seconds on a mid-tier computer and 2 minutes 4 seconds on a high-tier computer (See mid-tier and high-tier system configurations in Methods, Software graphical user interface development).

Video processing times are shown in **Supplementary eTable 4** and range from 5.7 to 16.6 times the video duration when tested on a mid-tier computer and 1.5 to 5.1 times the video duration when tested on a high-tier computer. Higher video resolution and frame rates increased processing times. Batch upload (vs. individual video upload) slightly reduced processing times.

## Discussion

We developed DxTonia as an open-source software package that detects leg dystonia in videos of children with CP performing a seated motor task. DxTonia achieves 88% sensitivity, 74% specificity, 82% PPV, 84% NPV, and 82% accuracy compared to gold standard expert consensus assessment during a seated hand open-close task. In contrast, we have shown that routine clinical evaluation for leg dystonia has 13% sensitivity, 11% specificity, 18% PPV, 8% NPV, and 12% accuracy for leg dystonia compared to gold standard expert consensus during gait.^13^ Because routine clinical care involves a single clinician assessing an individual with CP for dystonia in a time-limited fashion while also caring for their multiple other clinical needs, it is currently not feasible for a single clinician to achieve gold standard expert consensus diagnostic accuracy during routine clinical care, even if that clinician is an expert as we have previously shown.^13^ Even if clinical diagnostic accuracy for leg dystonia exceeds the 12% estimate based on gait assessment,^13^ the literature consistently shows that even expert clinicians have difficulty diagnosing dystonia.^50–52^ Our results suggest that DxTonia may facilitate screening for leg dystonia by analyzing brief videos acquired during routine clinical care. Our ultimate goal is for DxTonia to facilitate increased vigilance for dystonia in people with CP, thus triggering referrals for further dystonia evaluation, additional consideration of dystonia-targeted care, or more careful consideration of candidacy for spasticity-targeted surgical interventions.

In addition to developing DxTonia, we used conventional content analysis of expert consensus building discussions, ML methodologies, and calculation of feature importance using feature selection methods to identify 25 leg kinematic features likely to be most useful for detecting leg dystonia (**Table 2**). Features corresponding to variability of leg movements had higher importance rankings for detecting leg dystonia than did features corresponding to the amplitude of leg movements (**Table 2**, **Fig 3**). Therefore, though we previously identified that leg adduction variability and amplitude correlate with leg dystonia severity, our current results suggests that variable leg movement in multiple planes (not just leg adduction) enhance detection of leg dystonia. Therefore, independent of DxTonia, clinical vigilance for leg movement variability during a seated alternating hand open-close task could facilitate leg dystonia identification during routine clinical care.

### Limitations and areas for further optimization

We used the seated alternating hand open-close task for dystonia assessment. Though our previous work suggests that expert assessment of seated upper extremity tasks can approximate leg dystonia severity compared expert assessment of a comprehensive exam protocol,^23^ further work is necessary to directly compare leg dystonia assessment between the seated hand open-close task and gait. We also note that our metrics of clinical leg dystonia assessment accuracy are based on gait^13^ (routinely done in most movement-focused clinic visits), not on the seated alternating hand open-close task (which may be less routinely done). Future work should determine whether simply incorporating the seated hand open-close task into clinical assessment improves clinical leg dystonia assessment. Finally, though a seated task allows for dystonia assessment in people regardless of ambulation status, it still requires the cognitive ability to follow commands. Future work should assess how dystonia can be quantified outside of a standardized voluntary task.

Our study population was limited to people with CP. Though CP is the most common lifelong motor disability and the most common condition associated with dystonia in childhood,^2–4^ it is not the only condition associated with dystonia. Future work should determine whether DxTonia can detect leg dystonia due to other etiologies as well and whether tasks other than the seated alternating hand open-close task would be more useful to detect dystonia due to these other etiologies.

Video selection was crucial for model development and testing. The videos from Center 1 were shorter than videos from Center 2 as they only included one hand-open-close cycle, whereas Center 2 videos included five cycles. However, Center 1 had a larger number of videos than Center 2 (193 vs. 105). From a machine learning perspective, it is difficult to determine which dataset is preferable (longer videos or larger number). To minimize the effect of this data bias on model development and testing, and to allow for sufficient videos in the training, testing, and validation datasets, we selected videos from Center 1 for model development and testing, and videos from Center 2 for model testing only. Our use of five-fold cross-validation and five-fold testing also reduced sample biasing. The differences in videos between centers may have been a strength as DxTonia has now demonstrated encouraging results when tested on videos across two centers with different populations of people with CP, cameras, recording environments, and video durations, although these differences may have also contributed to differences in sensitivity and specificity for leg dystonia between these two centers. Therefore, testing DxTonia on additional datasets would further support its generalizability. In releasing DxTonia as an open-source tool, we encourage the ongoing optimization and validation of DxTonia across institutions and environmental conditions.

Though experts commonly cited toe movement features when assessing videos for leg dystonia (**Fig 2A),** it was difficult to reliably label multiple toes in our dataset using OpenPose. As open-source pose estimation continues to improve, we anticipate that incorporation of toe movement features may become more feasible and may further strengthen the diagnostic accuracy of DxTonia. Currently, DxTonia’s sensitivity is higher than its specificity for leg dystonia, supporting its utility as a screening tool. It is possible that inclusion of additional movement features may also improve DxTonia’s specificity for leg dystonia.

DxTonia is optimized to detect any dystonia (GDRS 1+). Dystonia rated as a GDRS of 1 may or may not have functional impact as the relationship between clinician-facing dystonia rating scales and functional impact remains unclear.^53^ Though a dystonia-specific functional impact scale has recently been validated in people with CP, it is caregiver-facing and thus requires significant education of the caregiver regarding how to differentiate dystonia from the myriad other movement manifestations of CP. Given these limitations, we were unable to identify what GDRS score corresponds to functional impact in our study population. However, we believe DxTonia is still useful as a leg dystonia screening tool noting that even mild leg dystonia can be useful to identify in young children with CP as this can dramatically affect counseling regarding surgical interventions.^7–10^ Still, it is possible that DxTonia is detecting dystonia that is subtle enough that it does not affect surgical outcomes or have functional impact. Future work should expand DxTonia to not only detect. Future work should expand DxTonia to not only detect dystonia but also to gauge its severity. Finally, noting that a standardized dystonia assessment would be valuable for clinical trials, future work should also assess the responsiveness of DxTonia to treatments.

Movement variability, though a key differentiator between dystonia and spasticity in this study, may also be a prominent feature of several other movement disorders, including ataxia, chorea, myoclonus, and tremor. DxTonia was developed specifically to differentiate dystonia from spasticity, by far the two most common, and typically co-occurring, movement symptoms in CP with prevalences far outpacing ataxia, chorea, myoclonus, and tremor in this population (which are collectively present in less than 3% of people with CP).^25^ Future work should focus on identifying quantifiable features differentiating dystonia from other movement disorders, not just spasticity.

All stages of leg dystonia assessment, other than data collection, are fully automated in the DxTonia software. Moving forward, we aim to develop an automated pipeline for data collection and secure local storage using smartphones, kinematics extraction via SecurePose^54^ (automated body pose estimation software), and generation of leg dystonia assessment reports. Such a system would improve the accessibility of leg dystonia assessment and facilitate widespread clinical implementation, optimization, and ongoing validation of DxTonia.

## Supporting information

Documentation for DxTonia

Best practice for recording the videos

Installed packages with their version in the anaconda virtual environment

Cross validation and testing

Software architecture of DxTonia

Demographic characteristics of children with CP in videos assessed for leg dystonia

Features and their importance rankings across eight feature selection methods averaged across all five dataset groupings

The CP characteristics of children with no spasticity or dystonia

DxTonia video processing time for different system configurations, video quality, and video upload methods

## Data Availability

All data produced in the present study are available upon reasonable request to the authors.

https://sourceforge.net/projects/dxtonia/

## Acknowledgement

This project is supported by Pediatric Epilepsy Research Foundation Registry Infrastructure Grant and the National Institute of Neurological Disorders and Stroke (NINDS 1K08NS117850-01A1). Scientific editing of this publication was supported by the Washington University Institute of Clinical and Translational Sciences grant UL1TR002345 from the National Center for Advancing Translational Sciences (NCATS) of the National Institutes of Health (NIH).

## Author Contributions

R. B. and B. A. contributed to the conception and design of the study; R. B., A. R., E.L., S. K., S. G., K. C., J. B., R. G. M, M. C. K., D. M., J. O. M., L. L. W., S. W., T. F. and B. A. contributed to the acquisition and analysis of data; R. B., J. S. P., and B. A. contributed to the drafting a significant portion of the manuscript or figures.

## Potential Conflicts of interest

The authors have no conflicts of interest to disclose.

## Data Availability

The videos used in the study are HIPPA protected data and cannot be shared. All other data, codes and the developed software are publicly available. Please refer to the project’s site (https://sourceforge.net/projects/dxtonia/).

## Notes

### Competing Interest Statement

The authors have declared no competing interest.

### Funding Statement

This study was funded by NINDS 1K08NS117850-01A1.

### Author Declarations

Institutional Review Board of Washington University in St. Louis gave ethical approval for this work (Approval Number: 202102101)

### Summary of Updates

Improved discussion section and overall readability. Added text is as follows “Even if clinical diagnostic accuracy for leg dystonia exceeds the 12% estimate based on gait assessment, the literature consistently shows that even expert clinicians have difficulty diagnosing dystonia.” "Still, it is possible that DxTonia is detecting dystonia that is subtle enough that it does not affect surgical outcomes or have functional impact" New supplementary material has also been added.

## References

1. Albanese, A. et al. Definition and Classification of Dystonia. Mov. Disord. 40, 1248–1259 (2025).

2. McIntyre, S. et al. Global prevalence of cerebral palsy: A systematic analysis. Dev. Med. Child Neurol. 64, 1494–1506 (2022).

3. Rice, J., Skuza, P., Baker, F., Russo, R. & Fehlings, D. Identification and measurement of dystonia in cerebral palsy. Dev. Med. Child Neurol. 59, 1249–1255 (2017).

4. Steeves, T. D., Day, L., Dykeman, J., Jette, N. & Pringsheim, T. The prevalence of primary dystonia: A systematic review and meta-analysis. Mov. Disord. 27, 1789–1796 (2012).

5. Fehlings, D. et al. Pharmacological and neurosurgical management of cerebral palsy and dystonia: Clinical practice guideline update. Dev. Med. Child Neurol. (2024) doi:10.1111/DMCN.15921.

6. Lott, E. et al. Physician Approaches to the Pharmacologic Treatment of Dystonia in Cerebral Palsy. Pediatrics 154, (2024).

7. Van De Pol, L. A. et al. Risk Factors for Dystonia after Selective Dorsal Rhizotomy in Nonwalking Children and Adolescents with Bilateral Spasticity. Neuropediatrics 49, 44–50 (2018).

8. Gillespie, C. S. et al. The effect of GMFCS level, age, sex, and dystonia on multi-dimensional outcomes after selective dorsal rhizotomy: prospective observational study. Childs. Nerv. Syst. 37, 1729–1740 (2021).

9. Blumetti, F. C. et al. Orthopaedic Surgery in Dystonic Cerebral Palsy. J. Pediatr. Orthop. 39, 209–216 (2019).

10. Buizer, A. I. et al. Effect of selective dorsal rhizotomy on daily care and comfort in non-walking children and adolescents with severe spasticity. Eur. J. Paediatr. Neurol. 21, 350–357 (2017).

11. Sanger, T. D., Delgado, M. R., Gaebler-Spira, D., Hallett, M. & Mink, J. W. Classification and Definition of Disorders Causing Hypertonia in Childhood. Pediatrics 111, e89–e97 (2003).

12. Kahlon, S. et al. Emerging Subspecialties: Pediatric Movement Disorders Neurology. Neurology 102, (2024).

13. Aravamuthan, B., Pearson, T. S., Chintalapati, K. & Ueda, K. Under-recognition of leg dystonia in people with cerebral palsy. Ann. Child Neurol. Soc. 1, 162–167 (2023).

14. Jethwa, A. et al. Development of the Hypertonia Assessment Tool (HAT): a discriminative tool for hypertonia in children. Wiley Online Libr. Jethwa, J Mink, C Macarthur, S Kn. T Fehlings, D FehlingsDevelopmental Med. Child Neurol. 2010•Wiley Online Libr. 52, (2010).

15. Knights, S. et al. Further evaluation of the scoring, reliability, and validity of the Hypertonia Assessment Tool (HAT). journals.sagepub.comS Kn. N Datoo, A Kawamura, L Switzer, D FehlingsJournal Child Neurol. 2014•journals.sagepub.com 29, 500–504 (2014).

16. Aravamuthan, B. R. et al. Gait features of dystonia in cerebral palsy. Dev. Med. Child Neurol. 63, 748–754 (2021).

17. Aravamuthan, B. R. et al. Determinants of gait dystonia severity in cerebral palsy. Dev. Med. Child Neurol. 65, 1–10 (2023).

18. Gemperli, K. et al. Chronic Striatal Cholinergic Interneuron Excitation Causes Cerebral Palsy-Related Dystonic Behavior in Mice. Ann. Neurol. (2025) doi:10.1002/ANA.27299.

19. Scott, I., Carter, S. & Coiera, E. Clinician checklist for assessing suitability of machine learning applications in healthcare. BMJ Heal. Care Informatics 28, e100251 (2021).

20. Supervised and Unsupervised Learning for Data Science. (2020) doi:10.1007/978-3-030-22475-2.

21. Salman, S. & Liu, X. Overfitting Mechanism and Avoidance in Deep Neural Networks. (2019).

22. Rosenbaum, P., Paneth, N., Leviton, A., Goldstein, M. & Bax, M. A report: the definition and classification of cerebral palsy April 2006. Dev. Med. Child Neurol. 49, 8–14 (2007).

23. Lott, E. et al. Tasks for assessing dystonia in young people with cerebral palsy. Ped Neurol 2025.09.07.25334597 at 10.1101/2025.09.07.25334597 (2025).

24. DxTonia download | SourceForge.net. https://sourceforge.net/projects/dxtonia/.

25. Kim, S. et al. Standardized clinical data capture to describe cerebral palsy. medRxiv [Preprint]. Ann Clin Trans Neurol [in Press] (2024) doi:10.1101/2024.08.09.24311474.

26. Comella, C. L., Leurgans, S., Wuu, J., Glenn, S. T. & Chmura, T. Rating scales for dystonia: a multicenter assessment. Mov. Disord. 18, 303–312 (2003).

27. Miles Matthew B, A. M. H. Qualitative Data Analysis. SAGE Publ. 1304, 354 (1994).

28. Cao, Z., Hidalgo, G., Simon, T., Wei, S. E. & Sheikh, Y. OpenPose: Realtime Multi-Person 2D Pose Estimation using Part Affinity Fields. IEEE Trans. Pattern Anal. Mach. Intell. 43, 172–186 (2018).

29. Cramer, J. S. The Origins of Logistic Regression. SSRN Electron. J. (2002) doi:10.2139/SSRN.360300.

30. Fürnkranz, J. Decision Tree. Encycl. Mach. Learn. 263–267 (2011) doi:10.1007/978-0-387-30164-8_204.

31. Fawagreh, K., Gaber, M. M. & Elyan, E. Random forests: from early developments to recent advancements. Syst. Sci. Control Eng. An Open Access J. 2, 602–609 (2014).

32. Bentéjac, C., Csörgő, A. & Martínez-Muñoz, G. A comparative analysis of gradient boosting algorithms. Artif. Intell. Rev. 54, 1937–1967 (2021).

33. Schapire, R. E. Explaining adaboost. Empir. Inference Festschrift Honor Vladimir N. Vapnik 37–52 (2013) doi:10.1007/978-3-642-41136-6_5/TABLES/2.

34. Schölkopf, B. SVMs - A practical consequence of learning theory. IEEE Intell. Syst. Their Appl. 13, 18–21 (1998).

35. Cunningham, P. & Delany, S. J. k-Nearest Neighbour Classifiers: 2nd Edition (with Python examples). ACM Comput. Surv. 54, (2020).

36. Vikramkumar, B, V. & Trilochan. Bayes and Naive Bayes Classifier. (2014).

37. Chen, T. & Guestrin, C. XGBoost: A Scalable Tree Boosting System. Proc. ACM SIGKDD Int. Conf. Knowl. Discov. Data Min. 13-17-August-2016, 785–794 (2016).

38. Geurts, P., Ernst, D. & Wehenkel, L. Extremely randomized trees. Mach. Learn. 63, 3–42 (2006).

39. BaggingClassifier — scikit-learn 1.5.1 documentation. https://scikit-learn.org/stable/modules/generated/sklearn.ensemble.BaggingClassifier.html.

40. Spearman Rank Correlation Coefficient. Concise Encycl. Stat. 502–505 (2008) doi:10.1007/978-0-387-32833-1_379.

41. Professor, M. S. G. Kendall’s Tau. Int. Encycl. Stat. Sci. 713–715 (2011) doi:10.1007/978-3-642-04898-2_324.

42. Theng, D. & Bhoyar, K. K. Feature selection techniques for machine learning: a survey of more than two decades of research. Knowl. Inf. Syst. 66, 1575–1637 (2024).

43. Zhao, Z., Anand, R. & Wang, M. Maximum Relevance and Minimum Redundancy Feature Selection Methods for a Marketing Machine Learning Platform. Proc. - 2019 IEEE Int. Conf. Data Sci. Adv. Anal. DSAA 2019 442–452 (2019) doi:10.1109/DSAA.2019.00059.

44. Aggrawal, R. & Pal, S. Sequential Feature Selection and Machine Learning Algorithm-Based Patient’s Death Events Prediction and Diagnosis in Heart Disease. SN Comput. Sci. 1, 1–16 (2020).

45. Guyon, I., Weston, J., Barnhill, S. & Vapnik, V. Gene selection for cancer classification using support vector machines. Mach. Learn. 46, 389–422 (2002).

46. Dietterich, T. G. Ensemble Methods in Machine Learning. Lect. Notes Comput. Sci. (including Subser. Lect. Notes Artif. Intell. Lect. Notes Bioinformatics) 1857 LNCS, 1–15 (2000).

47. ExifTool by Phil Harvey. https://www.exiftool.org/.

48. Lundh, F. An introduction to tkinter. URL www.pythonware.com/library/tkinter/introduction/index.htm (1999).

49. PyInstaller Manual — PyInstaller 6.16.0 documentation. https://pyinstaller.org/en/stable/.

50. Yilmaz, S., Vermilion, J., Dean, S., Pourdeyhimi, R. & Mink, J. W. Inter-rater Agreement for Movement Disorder Classification in Children with Hyperkinetic Movement Disorders. Mov. Disord. Clin. Pract. 11, 1598–1603 (2024).

51. Eggink, H. et al. Spasticity, dyskinesia and ataxia in cerebral palsy: Are we sure we can differentiate them? Eur. J. Paediatr. Neurol. 21, 703–706 (2017).

52. Beghi, E. et al. Reliability of clinical diagnosis of dystonia. Neuroepidemiology 43, 213– 219 (2014).

53. Stewart, K., … A. H.-D. M. &, 2017, undefined, Harvey, A. & Johnston, L. M. A systematic review of scales to measure dystonia and choreoathetosis in children with dyskinetic cerebral palsy. 59, 786–795 (2017).

54. Bajpai, R. & Aravamuthan, B. SecurePose: Automated Face Blurring and Human Movement Kinematics Extraction from Videos Recorded in Clinical Settings. arXiv (2025) 10.48550/arXiv.2402.14143.

