## Supplementary material for "Using expert-cited features to detect leg dystonia in cerebral palsy": Documentation for DxTonia

### Documentation for dxtonia.py

#### Overview

This is a Python script that creates a graphical user interface (GUI) for an application called OpenHKV: DxTonia. The GUI has several features, including:

1. **File Menu Bar**: A menu bar with options to:
    - Create a new project
    - Open an existing project
    - Select data
    - Save the project
    - Export patient data
    - Project information
    - Exit the application
2. **Kinematics Menu**: It allows users to perform various kinematic analysis tasks, including:
    - Video correction
    - Person detection
    - Person tracking
    - Smoothing tracking
    - Patient detection
3. **Dystonia Detection Menu**: It provides options for detecting dystonia in patients, including:
    - One-click dystonia detection
    - Leg dystonia detection (post-kinematic extraction)

The script uses the Tkinter library to create the GUI and its various components. It also imports several custom libraries, including `openhkv` and `splib`, which are specific to this application.

Some notable aspects of the code include:

- The use of global variables (`splib.global_var`) to store references to GUI frames and menus.
- The creation of a main window with a title "OpenHKV: DxTonia" and default size set to the full screen width and height.
- The use of event-driven programming, where the Tkinter event loop is started using `mainloop()`.

Overall, this script provides a comprehensive GUI for users to interact with the OpenHKV: DxTonia application, allowing them to perform various tasks related to kinematic analysis and dystonia detection.

### Documentation for splib:**init**.py

#### Overview

This file imports various pakages from library `splib`. Here's a summary of what each imported function does:

1. **`create_frame`**: This pakages is used to create new frames in a graphical user interface (GUI).
2. **`frame`**: This contains the definations of the frames and can be called from `create_frame` package.
3. **`sp`**: This pakage contains definations of all the core functions of the Python library `splib`.
4. **`global_var`**: As the name suggests, this handles global variables within the context of the `splib` library. This includes definiting and initiating the global variables values and system configurations.
5. **`window_setup`**: This pakage handelling GUI windows/frame, by setting up window-related properties, such as dimensions, title, or other visual attributes for a frame created using `create_frame`.

### Documentation for splib:create_frame.py

#### Overview

This packages is used to create new frames in a graphical user interface (GUI). Here's a summary of the code:

**Importing Libraries**

The script imports various libraries, including:

- `tkinter` for creating GUI windows
- `numpy`, `pandas`, and `scipy` for numerical computations
- `cv2` for computer vision operations
- `pickle` for data serialization and storage
- `json` for data formatting and storage
- and other libraries ...

**Clear screen**

The script destroys the objects of all frames. In other words, it closes all the frames.

**Create frames**

The script creates an object for a specific frame.

**Functions**

The script defines a series of functions, each of which creates or destroys a specific GUI frame. These functions include:

- `new_project_screen()`: Creates a new project screen
- `open_project_screen()`: Opens an existing project screen
- `select_data_screen()`: Creates a screen for selecting the data for processing
- `project_info_screen()`: Creates a screen to displays information about the current project
- `pre_process_screen()`: Creates a screen to preprocesses the videos
- `key_point_ext_screen()`: Creates a screen to extract the key points from the videos
- `set_ID_screen()`: Creates a screen to sets IDs for the person in each video of the project
- `intepolation_screen()`: Creates a screen to performs interpolation (find missing key point coordinates)
- `patient_detect_screen()`: Creates a screen to detects patients in the videos
- `bluring_screen()`: Creates a screen to blur the faces
- `bluring_quality_improvement_screen()`: Creates a screen to improve the blurring quality
- `oc_blurring_screen()`: Creates a screen to perform blurring in one-click
- `oc_dystonia_screen()`: Creates a screen to detect dystonia in one-click
- `dystonia_screen()`: Creates a screen to detect leg dystonia

Each function calls the `clear_screens()` function to destroy any existing GUI frames before creating a new one.

### Functions

#### clear_screens

- **Arguments**: None
- **Returns**: None
- **Description:**

The :func:`clear_screens` function is used to reset the GUI by destroying all frame objects that are currently visible. This function checks if each frame object exists and then destroys it if it does. If a frame object is not found, it simply skips over it.

This function should be called whenever a new project is created or an existing project is opened.

#### new_project_screen

- **Arguments**: None
- **Returns**: None
- **Description:**

Displays the new project screen, allowing users to create a new project.

Clears any existing screens, then instantiates a class:`NewProjectFrame` object and assigns it to the global variable `global_var.new_project_frame_obj`.

#### open_project_screen

- **Arguments**: None
- **Returns**: None
- **Description:**

This function is responsible for resetting the current screens and initializing the OpenProjectFrame object. It does not return any value, but rather sets the global variable `global_var.open_project_frame_obj` with the newly created frame object.

#### select_data_screen

- **Arguments**: None
- **Returns**: None
- **Description:**

Clears any existing screens, then instantiates a class:`SelectDataFrame` object and assigns it to the global variable `global_var.select_data_frame_obj`.

#### project_info_screen

- **Arguments**: None
- **Returns**: None
- **Description:**

Clears any existing screens, then instantiates a class:`ProjectInfoFrame` object and assigns it to the global variable `global_var.project_info_frame_obj`.

#### pre_process_screen

- **Arguments**: None
- **Returns**: None
- **Description:**

Clears any existing screens, then instantiates a class:`PreProcessFrame` object and assigns it to the global variable `global_var.pre_process_frame_obj`.

#### key_point_ext_screen

- **Arguments:** None
- **Returns**: None
- **Description:**

Clears any existing screens, then instantiates a class:`KeyPointExtFrame` object and assigns it to the global variable `global_var.key_point_ext_frame_obj`.

#### set_ID_screen

- **Arguments:** None
- **Returns**: None
- **Description:**

Clears any existing screens, then instantiates a class:`SetIDFrame` object and assigns it to the global variable `global_var.set_ID_frame_obj`.

#### intepolation_screen

- **Arguments:** None
- **Returns**: None
- **Description:**

Clears any existing screens, then instantiates a class:`IntepolationFrame` object and assigns it to the global variable `global_var.intepolation_frame_obj`.

#### patient_detect_screen

- **Arguments:** None
- **Returns**: None
- **Description:**

Clears any existing screens, then instantiates a class:`PatientDetectFrame` object and assigns it to the global variable `global_var.patient_detect_frame_obj`.

#### bluring_screen

- **Arguments:** None
- **Returns**: None
- **Description:**

Clears any existing screens, then instantiates a class:`BlurringFrame` object and assigns it to the global variable `global_var.blurring_frame_obj`.

#### bluring_quality_improvement_screen

- **Arguments:** None
- **Returns**: None
- **Description:**

Clears any existing screens, then instantiates a class:`BluringQualityImprovementFrame` object and assigns it to the global variable `global_var.bluring_quality_improvement_frame_obj`.

##### oc_blurring_screen

- **Arguments:** None
- **Returns**: None
- **Description:**

Clears any existing screens, then instantiates a class:`OCBlurringFrame` object and assigns it to the global variable `global_var.oc_blurring_frame_obj`.

##### oc_dystonia_screen

- **Arguments:** None
- **Returns**: None
- **Description:**

Clears any existing screens, then instantiates a class:`OCDystoniaFrame` object and assigns it to the global variable `global_var.oc_dystonia_frame_obj`.

##### dystonia_screen

- **Arguments:** None
- **Returns**: None
- **Description:**

Clears any existing screens, then instantiates a class:`DystoniaFrame` object and assigns it to the global variable `global_var.dystonia_frame_obj`.

### Documentation for splib:frame.py

#### Overview

This contains the definitions of the frames and can be called from `create_frame` package.

Properties of the each frame is represented by classes. The frame classes included in this package are:

- NewProjectFrame
- OpenProjectFrame
- SelectDataFrame
- ProjectInfoFrame
- PreProcessFrame
- KeyPointExtFrame
- SetIDFrame
- IntepolationFrame
- PatientDetectFrame

- BlurringFrame
- BluringQualityImprovementFrame
- SetupWindowFrame
- OCBlurringFrame
- OCDystoniaFrame
- DystoniaFrame

### Documentation for splib:global_var.py

#### Overview

As the name suggests, this handles global variables within the context of the `splib` library. This includes defining and initiating the global variables values and system configurations.

### Documentation for splib:sp.py

#### Overview

This package contains definitions of all the core functions of the Python library `splib`.

Here's a breakdown of the code:

**`oc_blurring()`**

1. Calls several functions to perform image processing:
    - `pre_process()`: Likely prepares the input data for further analysis.
    - `key_point_ext()`: Extracts key points from the processed data, possibly for feature extraction or tracking.
    - `set_ID()`: Sets an identifier (ID) for the current patient or subject.
    - `interpolation_face()`: Performs interpolation on the face data to create a more detailed representation.
    - `blurring()`: Applies blurring to the face data to reduce noise and improve analysis.
2. Enables the "Next" button in the GUI.

**`oc_dystonia()`**

1. Calls several functions to perform dystonia detection:
    - `pre_process()`: Prepares the input data for further analysis (similar to `oc_blurring()`).
    - `key_point_ext()`: Extracts key points from the processed data (similar to `oc_blurring()`).
    - `set_ID()`: Sets an identifier (ID) for the current patient or subject (similar to `oc_blurring()`).

  - `interpolation_leg()`: Performs interpolation on the leg data to create a more detailed representation.
  - `patient_detect()`: Detects dystonia in the patient based on the processed data.
2. Creates a GUI screen for displaying the dystonia detection results.

#### Functions

##### save_project

- **Arguments**: auto_save_flag: int Flag indicating whether to auto-save without prompt or prompt after manually saving. A value of 0 means manual saving, while a non-zero value enables auto-saving.
- **Return**: None
- **Description** :

  The function does not return any values. It saves the project data to disk and/or displays a confirmation message.

##### save_and_continue

- **Arguments**: None
- **Return**: None - The function does not return any value.
- **Description**

  Save the current project and continue with a clean slate. This function saves any unsaved changes to the current project, then clears all screens. It is typically used after completing an editing session or making significant changes to the project.

##### auto_save

- **Arguments**: None
- **Return**: None - The function does not return any value.
- **Description**

Automatically saves the current project.

##### create_project

- **Arguments**: None
- **Return**: None - The function does not return any value.
- **Description:**

This function is used to create a new project. It updates global variables to reflect the newly created project. It also updates the GUI to enable functionality related to the project,such as selecting data, saving the project, and exporting patient data. If an error occurs during directory creation (e.g., due to permission issues), an error message is displayed, and the log is updated with the error details. The function checks for the following user input: *`global_var.checkbutton_mute_value`: Value of the "Mute" check button * `global_var.project_name`: Name of the project entered by the user The function also updates the status and log accordingly.

#### open_project

- **Arguments**: None
- **Return**: None - The function does not return any value.
- **Description:**

Opens an existing project by loading its configuration files. The function retrieves the project name from the global variables, attempts to load the project's configuration files from disk, and updates the GUI accordingly. If successful, it enables various menu options and updates the application window title with the project name.

- **Raises**: OSError if the project cannot be loaded due to a disk error or other OS-related issue.

#### export_data

- **Arguments**: None
- **Return**: None - The function does not return any value.
- **Description:**

Exports video data from JSON files to CSV files.

This function iterates over each video, reads the corresponding JSON file, and extracts person coordinates. It then saves these coordinates to CSV files. The exported data is saved in the `coordinates` folder within the specified output path.

The function assumes that the global variables `global_var` are set and contain the necessary information for exporting data, including the working directory, project name, video names, and their corresponding JSON file paths.

#### load_videos

- **Arguments**: None

- **Return**: None - The function does not return any value.

- **Description:**

  Load video files from local system.

  Loads video files' information using a file dialog box. The selected videos' information are then stored in the global variables `video_path`, `video_name`, and video_paths_without_extension (their respective extensions removed). Finally, updates project status, log, and UI to reflect successful loading of videos.

The file dialog box allows selection of video files with the following extensions:

- .mp4 * .mov * .m4v

After loading, UI is updated to reflect project status and selected videos are made available for further processing.

#### load_meta

- **Arguments**: None
- **Return**: None - The function does not return any value.
- **Description:**

  Load meta data files from user-selected file paths. This function prompts the user to select meta data files using a file dialog. The selected file paths are stored in global variables for later use. Details of global variables modified:

data:`global_var.meta_path`: A list of file paths to selected meta data files. data:`global_var.meta_name`: A list of file names corresponding to the selected meta data files, without directory separators. data:`global_var.meta_names_without_extension`: A list of file names, minus their extensions (e.g. `file.txt` becomes `file`). data:`global_var.meta_paths_without_extension`: A list of file paths, minus their extensions (e.g. `/path/to/file.txt` becomes `/path/to/file`).

After loading the meta data files, this function updates the project status and saves any unsaved changes.

#### save_path

- **Arguments**: None
- **Return**: None - The function does not return any value.
- **Description:**

  Save path for data output folder and creates the folder. The function returns nothing (`None`) but updates several global variables with new paths and status information. These include:

---

```
-`global_var.data_save_path`: A list containing the selected directory path.
  - `global_var.new_video_path`: A list of video file paths with a generated ur
  - `global_var.new_json_path`: A list of JSON file paths with a generated uniq
```

The function attempts to create a new directory at the specified path. If successful, it updates the project status and shows a success message. If an error occurs (e.g., due to OS limitations), it displays an error message. Future version will have file name anonymization.

#### pre_process

- **Arguments**: None
- **Return**: None - The function does not return any value.
- **Description:**

Corrects video orientations using exiftool and saves the corrected videos. This function is responsible for correcting the orientations of the loaded videos. It uses `exiftool` to rotate each video based on its rotation angle and saves the corrected video in a new file.

The function follows these steps:

1. Initializes the logging and status updates by calling `window_setup.update_status` and `window_setup.update_log`.
2. Runs exiftool for each loaded video to extract its rotation angle and saves it in a text file.
3. Re-runs exiftool with the correct rotation angle to correct the orientation of each video.
4. Rotates each frame of the corrected videos using OpenCV's `cv2.rotate` function.
5. Saves the rotated frames as new videos in the output directory.

   The function updates the project status by setting `global_var.project_status[3] = 1`. It also calls the `auto_save` function to save the current state of the project.

   If all videos are corrected successfully, it enables the "Next" button and shows a success message. If any error occurs during the correction process, it displays an error message.

#### key_point_ext

- **Arguments**: None
- **Return**: None - The function does not return any value.
- **Description:**

  This function performs key point extraction using OpenPose on videos loaded by the application.

It updates the project status, displays a success message, and saves the extracted data to the designated folder.

This function is called after the video loading process has been completed. It assumes that the global variables `global_var.video_path`, `global_var.new_json_path`, `global_var.key_point_ext_frame_obj`, `global_var.project_status` are set correctly.

The function performs the following tasks:

1. Updates the status and log with a message indicating that key point extraction has started.
2. Changes the directory to the OpenPose build folder.
3. Iterates over each video in the loaded list, generating a command for OpenPose to perform key point extraction on each video.
4. Runs the generated commands using the `subprocess` module.
5. Updates the project status and saves the extracted data to the designated folder.

- **Exceptions** The function catches any OSError exceptions that may occur during execution, displaying an error message in a popup window and logging the error in the application log.

#### set_ID

- **Arguments**: None
- **Return**: None - The function does not return any value.
- **Description:**

The function assumes that the `all_video_json_path` attribute of the `global_var` object contains a list of paths to the video frames in JSON format. It also assumes that the `new_json_path` attribute of the `global_var` object is set. The function processes each video frame individually, assigns an ID to each person, and saves the updated frame as a JSON file. It logs progress and updates the project status.

#### interpolation_full

- **Arguments**: None
- **Return**: None - The function does not return any value.
- **Description:**

Smooths kinematics by interpolating missing frames of all key points. This function takes care of the interpolation process. It reads the json files, checks for bad frames, computes the weights, and finally replaces the coordinates of the person of interest in the original json files with the interpolated values.

Steps:

1. Reads JSON files from a specified directory and extracts keypoints for each frame.
2. Identifies bad frames based on keypoint data and stores them in a list.
3. Computes back index and weights, as well as forward index and weights, using bad frames data.
4. Interpolates keypoints at good frames by averaging values from neighboring frames weighted by their respective weights.
5. Writes interpolated keypoints to JSON files.

The following variables are used from the global namespace: - global_var.project_status[6] - global_var.video_name - global_var.video_names_without_extension - global_var.data_save_path -

global_var.window - global_var.intepolation_frame_obj.button_next['state']

After successful execution, this function updates the project status, logs a success message, and enables the 'Next' button.

- Raises: `OSError` if an error occurs during file I/O operations.

#### interpolation_face

- **Arguments**: None
- **Return**: None - The function does not return any value.
- **Description:**

Smooths kinematics by interpolating missing frames of the face key points. This function takes care of the interpolation process. It reads the json files, checks for bad frames, computes the weights, and finally replaces the coordinates of the person of interest in the original json files with the interpolated values.

Steps:

1. Reads JSON files from a specified directory and extracts keypoints for each frame.
2. Identifies bad frames based on keypoint data and stores them in a list.
3. Computes back index and weights, as well as forward index and weights, using bad frames data.
4. Interpolates keypoints at good frames by averaging values from neighboring frames weighted by their respective weights.
5. Writes interpolated keypoints to JSON files.

The following variables are used from the global namespace: - global_var.project_status[6] - global_var.video_name - global_var.video_names_without_extension - global_var.data_save_path - global_var.window - global_var.intepolation_frame_obj.button_next['state']

After successful execution, this function updates the project status, logs a success message, and enables the 'Next' button.

- Raises: `OSError` if an error occurs during file I/O operations.

#### interpolation_leg

- **Arguments**: None
- **Return**: None - The function does not return any value.
- **Description:**

Smooths kinematics by interpolating missing frames of the leg key points. This function takes care of the interpolation process. It reads the json files, checks for bad frames, computes the weights, and finally replaces the coordinates of the person of interest in the original json files with the interpolated values.

Steps:

1. Reads JSON files from a specified directory and extracts keypoints for each frame.
2. Identifies bad frames based on keypoint data and stores them in a list.
3. Computes back index and weights, as well as forward index and weights, using bad frames data.
4. Interpolates keypoints at good frames by averaging values from neighboring frames weighted by their respective weights.
5. Writes interpolated keypoints to JSON files.

The following variables are used from the global namespace: - global_var.project_status[6] - global_var.video_name - global_var.video_names_without_extension - global_var.data_save_path - global_var.window - global_var.intepolation_frame_obj.button_next['state']

After successful execution, this function updates the project status, logs a success message, and enables the 'Next' button.

- **Raises**: `OSError` if an error occurs during file I/O operations.

#### patient_detect

- **Arguments**: None
- **Return**: None - The function does not return any value.
- **Description:**

  This function identifies patients based on their gait patterns in videos. It uses OpenPose extracted keypoints from each video frame (stored in JSON files), then calculates distances between these keypoints to determine the unique identifier (set_ID function) of the patient. The ID close to the center of the video for the maximum duration is assigned as the patient ID.
- **Raises**: `OSError` if an error occurs during file I/O operations.

#### blurring

- **Arguments**: None
- **Return**: None - The function does not return any value.
- **Description:** This function processes all videos in the project folder. It reads video files, blurs faces in each frame of the video, and writes the blurred frames back to new video files. Face blurring uses a Gaussian blur with a kernel size of 99x99 pixels.
- **Raises**: `OSError` if an error occurs during file I/O operations.

#### manual_quality_improvement

- **Arguments**: None
- **Return**: None - The function does not return any value.

- **Description:** This function creates a Tkinter window with various features:
    - A slider for frame selection.
    - Sliders for selecting the start and end points of trimming.
    - Buttons to play, pause, and exit the video in play interval mode.
    - A button to edit the blurring. The function also calls other functions to create a timeline of video frames and bind events to Tkinter widgets.
- **Raises**: `OSError` if an error occurs during file I/O operations.

#### dystonia

- **Arguments**: None
- **Return**: None - The function does not return any value.
- **Description:** Main function for performing dystonia detection on video frames. This function performs the following steps:
    - Loads pre-trained models from file.
    - Prepares input data (video frames).
    - Extract features using `extract_features`
    - Performs predictions using loaded models.
    - Calculates final prediction by taking median of individual model predictions.
    - Writes ratings to CSV file for later analysis.
- **Raises**: `OSError` if an error occurs during file I/O operations.

#### extract_features

- **Arguments**: video_index : int The index of the video from which to extract features (0-based).
- **Return**: feature_mat : numpy.ndarray A 2D array where each row represents a video frame and each column represents a feature. The first column is reserved for future use.
- **Description:** Extracts features from a video at the specified index. This function loads the raw data of the video, computes various gait features, and returns them in a matrix format. The features extracted include standard deviations, minimum values, and maximum values of different joint coordinates.
- **Raises**: `OSError` if an error occurs during file I/O operations.

#### oc_blurring

- **Arguments**: None
- **Return**: None - The function does not return any value.
- **Description:**

  Perform face blurring in one-click by executing a series of steps.

This function performs a sequence of operations to blur faces detected in an input video frame, including pre-processing, key point extraction, ID setting, interpolation, and actual blurring. The result is stored for further processing or display.

##### oc_dystonia

- **Arguments**: None
- **Return**: None - The function does not return any value.
- **Description:**

  Performs leg dystonia detection in one-click by executing a series of steps including pre-processing, keypoint extraction, ID setting, interpolation, patient detection, and creating a dystonia screen.

### Documentation for splib:window_setup.py

#### Overview

###### Importing Libraries

The script imports various libraries, including:

- Tkinter for creating a graphical user interface (GUI)
- NumPy and Pandas for numerical computations
- OpenCV for image processing
- JSON and Pickle for data serialization
- Splib (SecurePose library) a custom library

###### Setup Window Function

The `setup_window()`:

1. Clears any existing screens using `splib.create_frame.clear_screens()`.
2. Creates a new window with a frame object, setting it as the global variable `global_var.setup_window_frame_obj`.
3. Initializes several global variables:
   - `project_status`: an array of 13 booleans representing the status of various tasks.
   - `working_directory`: the current working directory.

###### Update Project Status Function

The `update_project_status()` function updates the project status panel by:

1. Creating a string representation of the project status (e.g., "Pending" or "Completed").
2. Updating the text of the `label_project_status` widget in the GUI with the new status string.

**Update Status Functions**

Updates the status bar panel:

- `update_status(text, color=0)`: updates the status bar text and changes its color if specified.
- `update_log(text)`: appends a message to the log panel and writes it to a file in the project directory.

**Update Log Functions**

Updates the status bar panel and project log file:

- `update_status(text, color=0)`: updates the status bar text and changes its color if specified.
- `update_log(text)`: appends a message to the log panel and writes it to a file in the project directory.

### Functions

#### setup_window

- **Arguments:** None
- **Returns**: None
- **Description:**

The `setup_window()`:

1. Clears any existing screens using `splib.create_frame.clear_screens()`.
2. Creates a new window with a frame object, setting it as the global variable `global_var.setup_window_frame_obj`.
3. Initializes several global variables:
    - `project_status`: an array of 13 booleans representing the status of various tasks.
    - `working_directory`: the current working directory.

#### update_project_status

- **Arguments:** None
- **Returns**: None
- **Description:**

The `update_project_status()` function updates the project status panel by:

1. Creating a string representation of the project status (e.g., "Pending" or "Completed").

2. Updating the text of the `label_project_status` widget in the GUI with the new status string.

##### update_status

- **Arguments:**
    - **text** : `str`The text to be displayed on the status bar. This can be any string value.
    - **color** : `int` (optional)
        - A flag indicating whether to display the status in red or green color. If set to `0`, the default behavior is to display in green. Otherwise, it displays in red.
- **Returns**: None
- **Description:**

Updates the status bar panel:

- `update_status(text, color=0)`: updates the status bar text and changes its color if specified.
- `update_log(text)`: appends a message to the log panel and writes it to a file in the project directory.

##### update_log

- ***Arguments: text*** : *`str`The text to be displayed on the status bar. This can be any string value.*
- **Returns**: None
- **Description:**

Updates the status bar panel and project log file:

- `update_status(text, color=0)`: updates the status bar text and changes its color if specified.
- `update_log(text)`: appends a message to the log panel and writes it to a file in the project directory.

### Documentation for openhkv:**init**.py

#### Overview

The code is importing two modules from a library called openhkv:

1. `files`: This module contains functions or classes related to file operations, such as reading and writing files.
2. `gui`: This module contains functions or classes for creating graphical user interfaces (GUIs).

### Documentation for openhkv:files.py

#### Overview

Here is a summary of the code:

**Modules and Functions**

The code imports various modules, including:

- `tkinter` for GUI development
- `numpy` for numerical computations
- `json` and `pickle` for data serialization and deserialization
- `cv2` for image processing (not used in this code snippet)
- `pandas` for data manipulation (not used in this code snippet)

The code defines four main functions:

1. **save**: Saves a dictionary of values to a file using pickle.
2. **load**: Loads a dictionary from a file and sets global variables accordingly.
3. **read_json**: Reads a JSON file and returns its contents as a Python object.
4. **progressBar**: Displays a progress bar in the console. (only works with juputer-notebook)

**Global Variables**

The code uses global variables, which are accessed through the `globals()` function. The `save` and `load` functions modify these variables by adding or replacing values in the dictionary.

**Error Handling**

The code includes basic error handling for reading JSON files and loading data from pickle files.

#### Functions

##### save

- **Arguments**: filename (str), the name of the file to save to.
- **Description**:

  Save a dictionary of values to a binary file.

  This function takes a filename and any number of variable names as arguments. It creates a dictionary with these variables as keys, evaluates their current values using the `eval()` function, and saves this dictionary to the specified file in binary format using the `pickle` module.

##### load

- **Arguments**: filename, the path to the binary file containing the saved variables.

- **Description:** Loads variables from a binary file into the current global namespace. This function uses the exec() function to execute assignments in a loop, which can pose a security risk if the filename is not trusted.

##### read_json

- **Arguments**: file, the path to the JSON file to be read.

- **Return**: The parsed JSON data as a Python object, or None if an error occurs. rtype: Union[Dict, List, Int, Float, Bool, Str] or None

- **Description**:

  Reads and parses a JSON file into a Python object.

##### progressBar

- **Arguments**: count_value, total, suffix count_value (int): The current count or index of the ongoing task. total (int): The total number of items in the task. suffix (str, optional): A string to be appended at the end of the progress bar, indicating what has been completed so far. Defaults to an empty string.
- **Return**: None
- **Description**: Creates a progress bar in the console to display the progress of an ongoing task. This function prints a progress bar to the console based on the provided count value and total number of items. The progress bar is updated dynamically as the task progresses.

### Documentation for openhkv:gui.py

#### Overview

This code defines a class `ToolTip` and a function `CreateToolTip` that creates a tooltip for a given Tkinter widget. Here's how it works:

1. The `ToolTip` class is initialized with a Tkinter widget as an argument.
2. When the user hovers over the widget, the `showtip` method is called, which displays a tooltip window with the specified text at a position near the widget.
3. The `hidetip` method hides the tooltip window when the user moves away from the widget.
4. The `CreateToolTip` function takes two arguments: a Tkinter widget and some text to be displayed in the tooltip. It creates an instance of the `ToolTip` class, binds the `enter` and `leave` events to display/hide the tooltip accordingly.

**Key Features**

- Supports displaying tooltips for any Tkinter widget
- Customizable appearance (font, color, etc.) through label options
- Automatic hiding when user moves away from the widget

**Example Use Case**

```python
import tkinter as tk

root = tk.Tk()

button = tk.Button(root, text="Click me!")
CreateToolTip(button, "This is a tooltip")

root.mainloop()
```

In this example, a button with the label "Click me!" has a tooltip that displays when you hover over it.

### Functions

#### CreateToolTip

- **Arguments**: widget, text widget : :class:`tkinter.Widget` The widget that will have a tooltip displayed when hovered over. text : str The text to be displayed in the tooltip.
- **Description**:

Create a ToolTip widget to display text when hovering over another widget using `ToolTip` class.
