## Supplementary material for "Using expert-cited features to detect leg dystonia in cerebral palsy": Best practice for recording the videos

**eAppendix 1. Best practice for recording the videos**

**GENERAL GUIDELINES**

To ensure that video recordings capture patient movement with sufficient technical quality for accurate assessment by the proposed software, please follow these guidelines when setting up the recording device and preparing the patient:

- The camera should be elevated to the height of the patient’s abdominal region.
- The camera must remain in a fixed position (e.g., secured to a wall or stabilized with a tripod), not handheld. Also, it should be vertical to the ground and parallel to the patient’s frontal plane.
- The patient should be centered in the frame.
- Minimize the presence of other individuals in the video, as their inclusion may lead to inaccurate results.
- Avoid any interference, such as touching the patient’s body parts during recording.
- The patient’s bare limbs and bare feet should be visible. Avoid baggy clothing, such as long-sleeved shirts or loose pants, as this may lead to inaccurate results.
- Position the patient as close to the camera as possible while ensuring their entire body, from head to toe, remains fully visible in the frame throughout the video.
- Before uploading to the software, trim the video (only from start and end) to remove non-task-related segments, such as camera setup, instructions for the task, or unrelated movements by the patient.

**TASK PROTOCOLS**

**Seated Hand-Open-Close (HOC)**

**Setup**: For the seated hand-open-close (HOC) task, the patient should sit upright in a tall chair with proper back support, ensuring their feet do not touch the ground.

**Before starting the task**:

- The patient should be instructed to raise their dominant hand to face level, close their hand into a fist, then open it to familiarize themselves with the movement.
- The patient's hands should rest on their thighs, and their entire body, including their legs, should remain at rest.

**Task**:

- The patient begins with both palms on their thighs. They are instructed to raise their dominant hand to face level, close their hand into a fist, then open it to familiarize themselves with the movement.
- The patient should then open and close their hand 5 times as quickly as possible while maintaining control.
- During the task, the instructor should ensure that the patient fully opens and closes their hand, that their legs remain dangling in the air, and that the patient stays focused.
- After completing the task with their dominant hand, the patient should return both hands to their lap and remain still until any involuntary movements have stopped, or for two seconds.
- The task is then repeated with the non-dominant hand.
- The patient repeats the entire process two more times with each hand.
