## Supplementary material for "Using expert-cited features to detect leg dystonia in cerebral palsy": Installed packages with their version in the anaconda virtual environment

**eAppendix 2. Installed packages with their version in the anaconda virtual environment**

name: SECUREPOSE

channels:

- conda-forge

- anaconda

- defaults

dependencies:

| - _tflow_select=2.1.0 | - abseil-cpp=20210324.2 | - aiohttp=3.8.3 | - aiosignal=1.2.0 |
| --- | --- | --- | --- |
| - appdirs=1.4.4 | - astor=0.8.1 | - astunparse=1.6.3 | - async-timeout=4.0.2 |
| - attrs=22.1.0 | - blas=1.0 | - blinker=1.4 | - bottleneck=1.3.5 |
| - brotli-python=1.0.9 | - brotlipy=0.7.0 | - ca-certificates=2022.12.7 | - certifi=2022.12.7 |
| - cffi=1.15.1 | - chardet=5.1.0 | - click=8.0.4 | - colorama=0.4.6 |
| - cryptography=39.0.1 | - cudatoolkit=11.3.1 | - cudnn=8.2.1 | - dlib=19.24.0 |
| - flit-core=3.6.0 | - freeglut=3.2.2 | - freetype=2.10.4 | - frozenlist=1.3.3 |
| - gast=0.4.0 | - giflib=5.2.1 | - google-auth-oauthlib=0.4.6 | - google-pasta=0.2.0 |
| - h2=4.1.0 | - h5py=3.7.0 | - hdf5=1.10.6 | - hpack=4.0.0 |
| - hyperframe=6.0.1 | - icc_rt=2022.1.0 | - icu=68.1 | - idna=3.4 |
| - intel-openmp=2021.4.0 | - jasper=2.0.33 | - jpeg=9e | - keras-preprocessing=1.1.2 |
| - libblas=3.9.0 | - libcblas=3.9.0 | - libcurl=7.88.1 | - liblapack=3.9.0 |
| - liblapacke=3.9.0 | - libopencv=4.5.1 | - libpng=1.6.39 | - libprotobuf=3.17.2 |
| - libssh2=1.10.0 | - libtiff=4.2.0 | - libwebp-base=1.2.4 | - lz4-c=1.9.3 |
| - m2w64-gcc-libgfortran=5.3.0 | - m2w64-gcc-libs=5.3.0 | - m2w64-gcc-libs-core=5.3.0 | - m2w64-gmp=6.1.0 |
| - m2w64-libwinpthread-git=5.0.0.4634.697f757 | - markdown=3.4.1 | - mkl=2021.4.0 | - mkl-service=2.4.0 |
| - mkl_fft=1.3.1 | - mkl_random=1.2.2 | - msys2-conda-epoch=20160418 | - mtcnn=0.1.1 |
| - multidict=6.0.2 | - numexpr=2.8.4 | - oauthlib=3.2.2 | - opencv=4.5.1 |
| - openssl=1.1.1t | - opt_einsum=3.3.0 | - pandas=1.5.2 | - patsy=0.5.3 |
| - pip=23.0.1 | - pooch=1.4.0 | - py-opencv=4.5.1 | - pyasn1=0.4.8 |
| - pycparser=2.21 | - pyjwt=2.4.0 | - pyopenssl=23.0.0 | - pyreadline=2.1 |
| - pysocks=1.7.1 | - python=3.9.16 | - python-dateutil=2.8.2 | - python_abi=3.9 |
| - pytz=2022.7 | - pyu2f=0.1.5 | - pyyaml=6.0 | - qt=5.12.9 |
| - rsa=4.9 | - scipy=1.10.0 | - setuptools=65.6.3 | - six=1.16.0 |
| - snappy=1.1.9 | - sqlite=3.40.1 | - statsmodels=0.13.5 | - tensorboard-data-server=0.6.1 |
| - tensorboard-plugin-wit=1.8.1 | - tensorflow=2.6.0 | - tensorflow-base=2.6.0 | - tensorflow-gpu=2.6.0 |
| - vc=14.2 | - vs2015_runtime=14.27.29016 | - wheel=0.35.1 | - win_inet_pton=1.1.0 |
| - wincertstore=0.2 | - xz=5.2.6 | - yaml=0.2.5 | - yarl=1.8.1 |
| - zlib=1.2.13 | - zstandard=0.18.0 | - zstd=1.5.0 |  |

- pip:

| - absl-py==1.4.0 | - altgraph==0.17.3 | - anyio==3.6.2 | - argon2-cffi==21.3.0 |
| --- | --- | --- | --- |
| - argon2-cffi-bindings==21.2.0 | - arrow==1.2.3 | - asttokens==2.2.1 | - backcall==0.2.0 |
| - beautifulsoup4==4.12.2 | - bleach==6.0.0 | - cachetools==5.3.0 | - category-encoders==2.6.0 |
| - charset-normalizer==3.1.0 | - comm==0.1.3 | - contourpy==1.0.7 | - cycler==0.11.0 |
| - cython==0.29.34 | - debugpy==1.6.7 | - decorator==5.1.1 | - defusedxml==0.7.1 |
| - docopt==0.6.2 | - et-xmlfile==1.1.0 | - executing==1.2.0 | - fastjsonschema==2.16.3 |
| - flatbuffers==23.3.3 | - fonttools==4.39.3 | - fqdn==1.5.1 | - google-auth==2.16.2 |
| - grpcio==1.51.3 | - importlib-metadata==6.0.0 | - importlib-resources==5.12.0 | - ipykernel==6.22.0 |
| - ipython==8.12.0 | - ipython-genutils==0.2.0 | - ipywidgets==8.0.6 | - isoduration==20.11.0 |
| - jedi==0.18.2 | - jinja2==3.1.2 | - joblib==1.2.0 | - js2py==0.74 |
| - jsonpointer==2.3 | - jsonschema==4.17.3 | - jupyter==1.0.0 | - jupyter-client==8.1.0 |
| - jupyter-console==6.6.3 | - jupyter-core==5.3.0 | - jupyter-events==0.6.3 | - jupyter-server==2.5.0 |
| - jupyter-server-terminals==0.4.4 | - jupyterlab-pygments==0.2.2 | - jupyterlab-widgets==3.0.7 | - keras==2.11.0 |
| - kiwisolver==1.4.4 | - libclang==15.0.6.1 | - markupsafe==2.1.2 | - matplotlib==3.7.1 |
| - matplotlib-inline==0.1.6 | - mistune==2.0.5 | - mlxtend==0.22.0 | - mrmr-selection==0.2.6 |
| - nbclassic==0.5.5 | - nbclient==0.7.3 | - nbconvert==7.3.0 | - nbformat==5.8.0 |
| - nest-asyncio==1.5.6 | - notebook==6.5.4 | - notebook-shim==0.2.2 | - numpy==1.24.2 |
| - openpyxl==3.1.2 | - packaging==23.0 | - pandocfilters==1.5.0 | - parso==0.8.3 |
| - pefile==2023.2.7 | - pickleshare==0.7.5 | - pillow==9.5.0 | - pipwin==0.5.2 |
| - platformdirs==3.2.0 | - prometheus-client==0.16.0 | - prompt-toolkit==3.0.38 | - protobuf==3.19.6 |
| - psutil==5.9.4 | - pure-eval==0.2.2 | - pyasn1-modules==0.2.8 | - pydot==1.4.2 |
| - pydotplus==2.0.2 | - pygments==2.14.0 | - pyinstaller==6.10.0 | - pyinstaller-hooks-contrib==2024.8 |
| - pyjsparser==2.7.1 | - pyparsing==3.0.9 | - pyprind==2.11.3 | - pyrsistent==0.19.3 |
| - pysmartdl==1.3.4 | - python-json-logger==2.0.7 | - python-version==0.0.2 | - pywin32==306 |
| - pywin32-ctypes==0.2.3 | - pywinpty==2.0.10 | - pyzmq==25.0.2 | - qtconsole==5.4.2 |
| - qtpy==2.3.1 | - requests==2.28.2 | - requests-oauthlib==1.3.1 | - rfc3339-validator==0.1.4 |
| - rfc3986-validator==0.1.1 | - scikit-learn==1.2.2 | - send2trash==1.8.0 | - sniffio==1.3.0 |
| - soupsieve==2.4 | - stack-data==0.6.2 | - tensorboard==2.11.2 | - tensorflow-estimator==2.11.0 |
| - tensorflow-intel==2.11.0 | - tensorflow-io-gcs-filesystem==0.31.0 | - termcolor==2.2.0 | - terminado==0.17.1 |
| - threadpoolctl==3.1.0 | - tinycss2==1.2.1 | - tornado==6.2 | - tqdm==4.65.0 |
| - traitlets==5.9.0 | - typing-extensions==4.5.0 | - tzdata==2024.1 | - tzlocal==5.2 |
| - uri-template==1.2.0 | - urllib3==1.26.15 | - wcwidth==0.2.6 | - webcolors==1.13 |
| - webencodings==0.5.1 | - websocket-client==1.5.1 | - werkzeug==2.2.3 | - widgetsnbextension==4.0.7 |
| - wrapt==1.15.0 | - xgboost==2.0.3 | - zipp==3.15.0 |  |

Custom libraries

- openhkv

- splib
