## Supplementary material for "Using expert-cited features to detect leg dystonia in cerebral palsy": Cross validation and testing

**eFigure 1. Cross validation and testing.** The data from Center 1 was used to create five dataset groupings with different training and testing sets. Then, we used a grid search methodology to conduct feature selection and model development and identify the best performing model and their corresponding feature sets for each dataset grouping. Finally, we merged all five best performing models using an average ensemble technique to obtain a final prediction. These models were then tested on the dataset from Center 2.

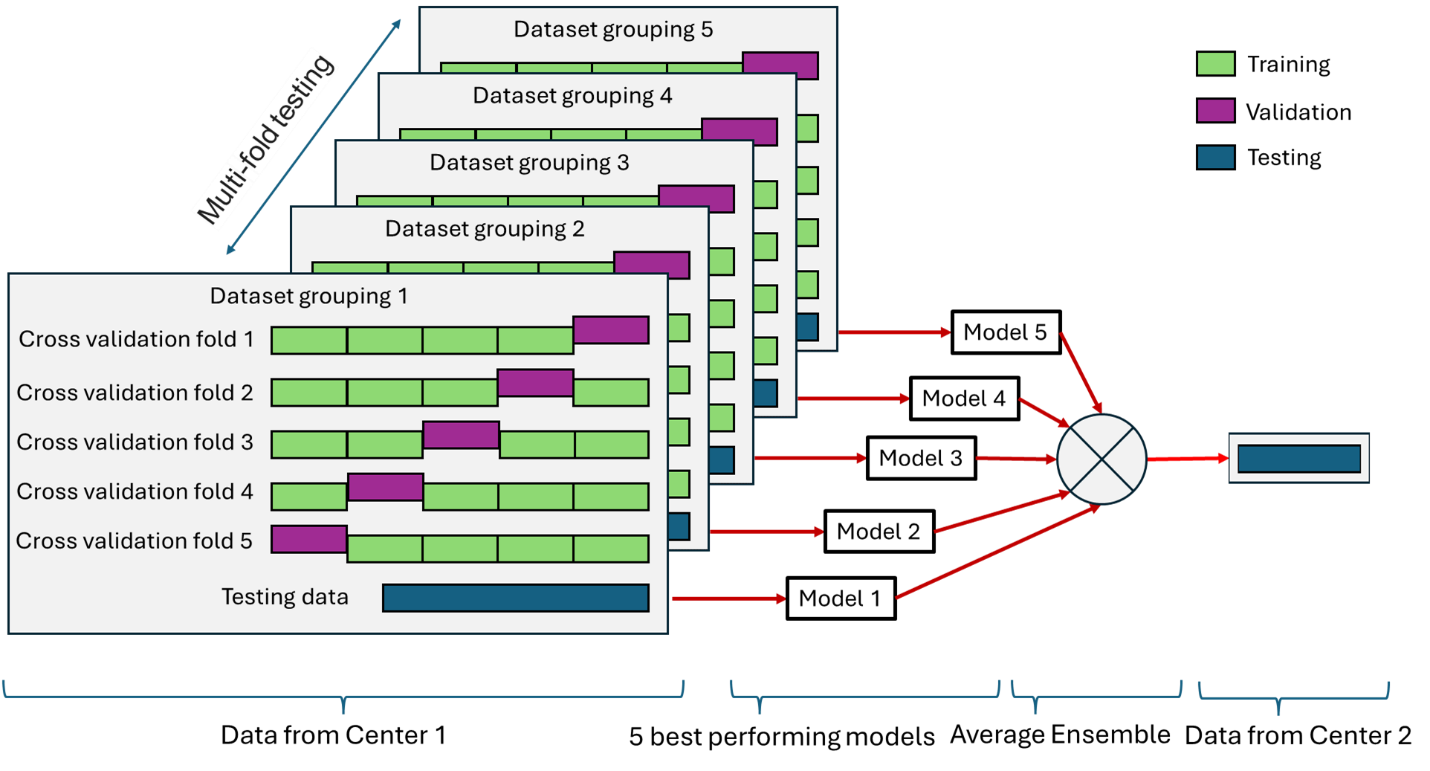
