## Supplementary material for "Using expert-cited features to detect leg dystonia in cerebral palsy": Software architecture of DxTonia

**eFigure 2. Software architecture of DxTonia:** The illustration shows modules used by DxTonia for the video processing and leg dystonia detection. Code and the software are freely available at <https://sourceforge.net/projects/dxtonia/> .

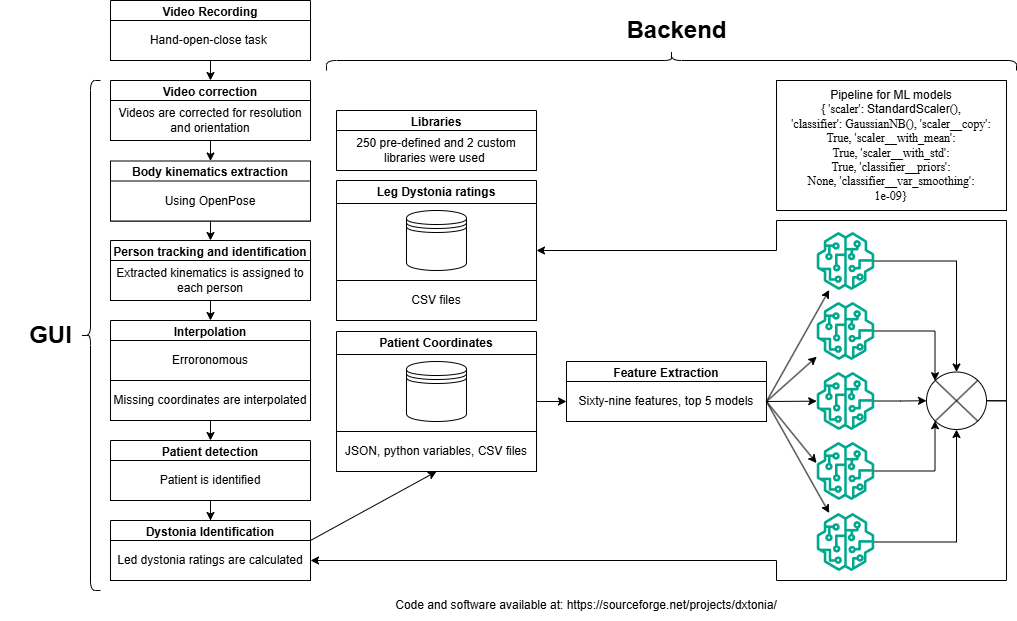
