## Supplementary material for "Using expert-cited features to detect leg dystonia in cerebral palsy": Demographic characteristics of children with CP in videos assessed for leg dystonia

**eTable 1. Demographic characteristics of children with CP in videos assessed for leg dystonia.**

|  | **Center 1** | | | **Center 2** | | |
| --- | --- | --- | --- | --- | --- | --- |
|  | No dystonia  (GDRS < 1),  N = 89 | Dystonia  (GDRS ≥ 1),  N = 104 | Missing Data  (n) | No dystonia  (GDRS < 1),  N = 42 | Dystonia  (GDRS ≥ 1),  N = 63 | Missing Data  (n) |
| Age (years, mean ± standard deviation) | 7.6 ± 3.3 | 6.9 ±2.9 | 6 | 11.7±3.4 | 10.4±3.8 |  |
| Gestational age (weeks, mean ± standard deviation) | 32.3 ± 6.4 | 31.6 ± 5.6 | 9 | 33±6.6 | 35 ±5.7 | 9 |
| CP Distribution (n, %) |  |  | 10 |  |  | 1 |
| Hemiplegia | 7, 8% | 7, 7% |  | 13, 31% | 32, 52% |  |
| Diplegia | 66, 78% | 53, 55% |  | 18, 43% | 15, 24% |  |
| Triplegia | 9, 11% | 21, 22% |  | 2, 5% | 5, 8% |  |
| Quadriplegia | 3, 4% | 15, 16% |  | 7, 17% | 9, 15% |  |
| none | 3, 4% | 3, 4% |  | 2, 5% | 1, 2% |  |
| GMFCS (n, %) |  |  | 17 |  |  |  |
| I | 17, 22% | 18, 19% |  | 19, 45% | 25, 40% |  |
| II | 36, 47% | 36, 37% |  | 19, 45% | 35, 56% |  |
| III | 19, 25% | 33, 34% |  | 4, 10% | 3, 5% |  |
| IV | 4, 5% | 10, 10% |  | 0, 0% | 0, 0% |  |
| V | 0, 0% | 0, 0% |  | 0, 0% | 0, 0% |  |
| Leg spasticity (n, %) |  |  | 6 | Spasticity (n, %) | |  |
| Spasticity absent | 7, 8% | 9, 10% |  | 7, 17% | 7, 11% |  |
| Spasticity present | 80, 92% | 91, 91% |  | 35, 83% | 56, 89% |  |
| GDRS – Global dystonia rating scale. GMFCS – Gross Motor Functional Classification System Level (I-II – independently ambulatory without assistive devices, III – independently ambulatory with assistive devices, IV-V – predominantly uses wheelchair for mobility). | | | | | | |
