## Supplementary material for "Using expert-cited features to detect leg dystonia in cerebral palsy": Features and their importance rankings across eight feature selection methods averaged across all five dataset groupings

**eTable 2. Features and their importance rankings across eight feature selection methods averaged across all five dataset groupings. |x| represents absolute value of x.**

| Feature Importance Rank | Features |
| --- | --- |
| 1 | Variance \| Left Toe (X) – Left Ankle (X) \| |
| 2 | Variance \|Right Toe (X) – Right Ankle (X)\| |
| 3 | Variance \| Left Toe (X) – Right Toe (X) \| |
| 4 | Variance Left Toe (Y) |
| 5 | Variance \| Left Ankle (X) – Right Ankle (X) \| |
| 6 | Variance Left Toe (X) |
| 7 | Variance Right Toe (X) |
| 8 | Variance Right Toe (Y) |
| 9 | Variance Right Ankle (Y) |
| 10 | Variance Right Ankle (X) |
| 11 | Variance \| Left Toe (Y) – Left Ankle (Y) \| |
| 12 | Variance \| Left Toe (Y) – Left Knee (Y) \| |
| 13 | Variance Left Ankle (Y) |
| 14 | Variance Left Ankle (X) |
| 15 | Variance \| Right Toe (Y) – Right Ankle (Y) \| |
| 16 | Maximum \| Left Toe (Y) – Left Ankle (Y) \| |
| 17 | Minimum \| Left Toe (Y) – Left Ankle (Y)\| |
| 18 | Maximum \| Right Toe (Y) – Right Ankle (Y) \| |
| 19 | Minimum \| Right Toe (Y) – Right Ankle (Y) \| |
| 20 | Variance \| Left Toe (X) - Left Knee (X) \| |
| 21 | Variance \| Right Toe (Y) – Right Knee (Y) \| |
| 22 | Variance \| Right Toe (X) – Right Knee (X) \| |
| 23 | Minimum Left Toe (Y) |
| 24 | Maximum \| Left Ankle (X) – Right Ankle (X) \| |
| 25 | Maximum Left Toe (Y) |
| 26 | Maximum Right Toe (Y) |
| 27 | Minimum \| Left Ankle (X) – Right Ankle (X) \| |
| 28 | Maximum Left Toe (X) |
| 29 | Maximum \| Left Toe (X) – Right Toe (X) \| |
| 30 | Minimum \| Left Knee (X) – Right Knee (X) \| |
| 31 | Minimum \| Left Toe (X) – Right Toe (X) \| |
| 32 | Minimum Right Toe (Y) |
| 33 | Variance \| Left Knee (X) – Right Knee (X) \| |
| 34 | Variance Left Knee (X) |
| 35 | Minimum Left Toe (X) |
| 36 | Maximum \| Left Toe (Y) – Left Knee (Y) \| |
| 37 | Maximum \| Left Knee (X) – Right Knee (X) \| |
| 38 | Maximum Right Toe (X) |
| 39 | Minimum \| Right Toe (Y) – Right Knee (Y) \| |
| 40 | Maximum \| Right Toe (Y) – Right Knee (Y) \| |
| 41 | Minimum \| Left Toe (Y) – Left Knee (Y) \| |
| 42 | Variance Right Knee (X) |
| 43 | Maximum Left Knee (X) |
| 44 | Maximum Left Ankle (Y) |
| 45 | Minimum Right Ankle (Y) |
| 46 | Maximum Right Ankle (Y) |
| 47 | Minimum Left Knee (X) |
| 48 | Variance Right Knee (Y) |
| 49 | Minimum Left Ankle (Y) |
| 50 | Minimum Right Toe (X) |
| 51 | Variance Left Knee (Y) |
| 52 | Maximum Left Ankle (X) |
| 53 | Minimum Left Ankle (X) |
| 54 | Minimum Left Knee (Y) |
| 55 | Maximum Left Knee (Y) |
| 56 | Maximum \| Right Toe (X) – Right Ankle (X) \| |
| 57 | Minimum Right Knee (X) |
| 58 | Minimum Right Ankle (X) |
| 59 | Maximum Right Ankle (X) |
| 60 | Maximum Right Knee (X) |
| 61 | Minimum Right Knee (Y) |
| 62 | Minimum \| Right Toe (X) – Right Ankle (X) \| |
| 63 | Maximum Right Knee (Y) |
| 64 | Maximum \| Left Toe (X) – Left Knee (X) \| |
| 65 | Maximum \| Left Toe (X) – Left Ankle (X) \| |
| 66 | Minimum \| Left Toe (X) – Left Ankle (X) \| |
| 67 | Minimum \| Left Toe (X) – Left Knee (X) \| |
| 68 | Maximum \| Right Toe (X) – Right Knee (X) \| |
| 69 | Minimum \| Right Toe (X) – Right Knee (X) \| |
