## Supplementary material for "Using expert-cited features to detect leg dystonia in cerebral palsy": The CP characteristics of children with no spasticity or dystonia

**eTable 3. The CP characteristics of children with no spasticity or dystonia.**

|  | **Center 1** | **Center 2** |
| --- | --- | --- |
| **Number of children** | N = 7 | N = 7 |
| **Age (years, mean ± standard deviation)** | 7.24 ± 2.8 | 10.95 ± 4.3 |
| **CP Distribution (n, %)** |  |  |
| **Hemiplegia** | 1, 14% | 0, 0% |
| **Diplegia** | 5, 72% | 0, 0% |
| **Triplegia** | 0, 0% | 0, % |
| **Quadriplegia** | 1, 14% | 6, 86% |
| **none** | 0, 0% | 1, 14% |
| **GMFCS (n, %)** |  |  |
| **I** | 3, 43% | 1, 14% |
| **II** | 1, 14% | 5, 72% |
| **III** | 3, 43% | 0, 0% |
| **IV** | 0, 0% | 0, 0% |
| **V** | 0, 0% | 1, 14% |
