## Supplementary material for "Using expert-cited features to detect leg dystonia in cerebral palsy": DxTonia video processing time for different system configurations, video quality, and video upload methods

**eTable 4.** **DxTonia video processing time for different system configurations, video quality, and video upload methods.**

| **For a high-tier consumer computer: IntelR Core™ i9 13900K CPU @ 3.00 GHz 24 cores, 196-GB RAM, NVIDIA GeForce RTX4090 GPU with 24GB memory, 64-bit Windows 11 Pro Operating System** | | | | | |
| --- | --- | --- | --- | --- | --- |
| **Video upload method (individual vs. batch upload of 10 videos) and video quality (low vs. high)** | **Video Resolution** | **Video fps** | **Video Duration**  **(mean±sd)** | **Processing time**  **(mean±sd)** | **Processing time to duration ratio** |
| Individual upload, Low quality | 1280×720 pixels | 30 | 5.8±2.2 seconds | 12±6 seconds | 2 |
| Individual upload, High quality | 3840×2160 pixels | 60 | 67±18 seconds | 340±99 seconds | 5.1 |
| Batch upload, Low quality | 1280×720 pixels | 30 | 5.8±2.2 seconds | 9±4  seconds | 1.5 |
| Batch upload, High quality | 3840×2160 pixels | 60 | 67±18 seconds | 321±97 seconds | 4.8 |
| **For a mid-tier consumer computer: IntelR Core™ i7 11800H CPU @ 2.40 GHz 16 cores, 32-GB RAM, NVIDIA GeForce RTX3070 GPU with 8GB memory, 64-bit Windows 11 Pro Operating System** | | | | | |
| **Video upload method (individual vs. batch upload of 10 videos) and video quality (low vs. high)** | **Video Resolution** | **Video fps** | **Video Duration (mean±sd)** | **Processing time**  **(mean±sd)** | **Processing time to duration ratio** |
| Individual upload, Low quality | 1280×720 pixels | 30 | 5.8±2.2 seconds | 38±16 seconds | 6.5 |
| Individual upload, High quality | 3840×2160 pixels | 60 | 67±18 seconds | 1112±256 seconds | 16.6 |
| Batch upload, Low quality | 1280×720 pixels | 30 | 5.8±2.2 seconds | 33±10 seconds | 5.7 |
| Batch upload, High quality | 3840×2160 pixels | 60 | 67±18 seconds | 999±255 seconds | 14.9 |
| SD: standard deviation, Low quality video: 1280×720 pixels, 30 fps, High quality video: 3840×2160 pixels, 60 fps. | | | | | |
